# Lasting first impression: Pre-existing immunity restricts mucosal antibody responses during Omicron breakthrough

**DOI:** 10.1101/2023.03.28.23287848

**Authors:** Kevin John Selva, Pradhipa Ramanathan, Ebene Regina Haycroft, Arnold Reynaldi, Deborah Cromer, Chee Wah Tan, Lin-Fa Wang, Bruce D Wines, P Mark Hogarth, Laura E Downie, Samantha K Davis, Ruth Amy Purcell, Helen E Kent, Jennifer A Juno, Adam K Wheatley, Miles P Davenport, Stephen John Kent, Amy W Chung

**Affiliations:** Department of Microbiology and Immunology, Peter Doherty Institute for Infection and Immunity, University of Melbourne, Melbourne, Victoria, 3000, Australia; Kirby Institute, University of New South Wales, Kensington, New South Wales, 2052, Australia; Programme in Emerging Infectious Diseases, Duke-NUS Medical School, 169857, Singapore; Singhealth Duke-NUS Global Health Institute, 169856, Singapore; Immune Therapies Laboratory, Burnet Institute, Melbourne, Victoria, 3004 Australia; Department of Immunology and Pathology, Central Clinical School, Monash University, Melbourne, Victoria, 3800, Australia; Department of Clinical Pathology, University of Melbourne, Melbourne, Victoria, 3000, Australia; Department of Optometry and Vision Sciences, University of Melbourne, Carlton, Victoria, 3053, Australia; Melbourne Sexual Health Centre and Department of Infectious Diseases, Alfred Hospital and Central Clinical School, Monash University, Melbourne, VIC, Australia

**Keywords:** COVID-19, mRNA vaccine, mucosal antibodies, saliva, breakthrough infection, Omicron, IgG4, Fc receptor, pre-existing immunity, IgA

## Abstract

Understanding mucosal antibody responses from SARS-CoV-2 infection and/or vaccination is crucial to develop strategies for longer term immunity, especially against emerging viral variants. We profiled serial paired mucosal and plasma antibodies from: COVID-19 vaccinated only vaccinees (vaccinated, uninfected), COVID-19 recovered vaccinees (convalescent, vaccinated) and individuals with breakthrough Delta or Omicron BA.2 infections (vaccinated, infected). Saliva from COVID-19 recovered vaccinees displayed improved antibody neutralizing activity, FcγR engagement and IgA compared to COVID-19 uninfected vaccinees. Furthermore, repeated mRNA vaccination boosted SARS-CoV-2-specific IgG2 and IgG4 responses in both mucosa biofluids (saliva and tears) and plasma. IgG, but not IgA, responses to breakthrough COVID-19 variants were dampened and narrowed by increased pre-existing vaccine-induced immunity to the ancestral strain. Salivary antibodies delayed initiation of boosting following breakthrough COVID-19 infection, especially Omicron BA.2, however, rose rapidly thereafter. Our data highlight how pre-existing immunity shapes mucosal SARS-CoV-2-specific antibody responses and has implications for long-term protection from COVID-19.

## Introduction

COVID-19 vaccines, and particularly mRNA boosters, elicit SARS-CoV-2 specific neutralizing antibodies systemically and protect against severe disease. However, current intramuscular (IM) COVID-19 vaccination regimes among SARS-CoV-2 uninfected individuals induce limited site-specific neutralizing antibodies at the mucosa – the site of SARS-CoV-2 acquisition [1]. This gap in mucosal humoral immunity is thought to contribute to vaccine breakthrough infections [2, 3].

Prior SARS-CoV-2 infection primes improved mucosal antibody responses elicited by subsequent vaccinations [2, 4-6]. Previous studies have largely focused on the neutralizing potential of mucosal antibody isotypes IgG and IgA, with little known about mucosal antibody subclass responses (IgG1-4, IgA1-2), each of which have unique profiles and functions. Furthermore, while the retention of antibody-mediated functional responses, despite the waning of neutralization, has been demonstrated in the blood, its potential at the mucosal surface remains understudied [7].

Pre-existing vaccine-induced immunity may also modulate immune responses during breakthrough infections. Breakthrough infections with the more divergent Omicron BA.1 strain is associated with a more modest recall of SARS-CoV-2 spike immunity as compared to Delta breakthroughs [8]. Immunological imprinting from repeated vaccinations with the ancestral spike may hamper the development of robust systemic humoral responses specific against Omicron during breakthrough infections [9-11]. Unfortunately, most studies have only focused on systemic antibodies, and the impact of prior ancestral strain vaccination on mucosal antibodies following breakthrough infection is unclear.

Herein, we compare COVID-19 recovered (convalescent, vaccinated) and vaccinated only vaccinees (vaccinated, uninfected), demonstrating that recovered individuals elicit stronger mucosal antibodies following vaccination, with higher capacity to induce antibody-mediated functional responses. Furthermore, using a series of paired mucosal and plasma samples collected very early following Delta and Omicron BA.2 breakthrough infections (vaccinated then infected), we demonstrate that pre-existing immunity also differentially impacts mucosal immunity.

## Materials and Methods

### Cohort and sample collection

We enrolled individuals with and without prior SARS-CoV-2 infection from a previously described cohort [12] to donate blood and saliva prior to and following vaccinations with either BNT162b2 (Comirnaty; Pfizer-BioNTech) or ChAdOx1 nCoV-19 (AstraZeneca) vaccines, as well as mRNA boosters (Supp Figure 1a).

We also recruited previously vaccinated individuals with a nasal PCR-confirmed breakthrough COVID-19 [8, 13, 14] during the Delta and Omicron BA.2 waves in Victoria to provide serial blood and saliva samples (Supp Figure 1b, c).

Whole blood was collected with sodium heparin anticoagulant and plasma was collected and stored at -80°C until use. Saliva was collected by SalivaBio Oral Swabs (Salimetrics) and processed following manufacturer’s instructions, before being stored at -80°C until use. Basal (non-stimulated) tear samples (∼7□μL per eye) were collected by capillary flow (Drummond Scientific, Broomall, PA, USA) from the inferior tear meniscus as previously reported, and also stored at -80°C until use [15].

### Ethics

The study protocols were approved by the University of Melbourne Human Research Ethics Committee (2021-21198-15398-3, 2056689, 11507), and all associated procedures were carried out in accordance with approved guidelines. All participants provided written informed consent in accordance with the Declaration of Helsinki.

### Ancestral SARS-CoV-2 multiplex bead assay

SARS-CoV-2 specific antibody isotypes (IgG, IgA, IgM) and subclasses (IgG1-4, IgA1-2) in plasma (1:1600), saliva (1:12.5) and tear (1:25) from the respective pre-pandemic and vaccinated cohorts were assessed using a customized multiplex bead-based array consisting of 4 ancestral SARs-CoV-2 proteins, including whole Spike Trimer (ST), Spike 1 (S1), Spike 2 (S2) and Receptor Binding Domain (RBD) as previously described [1] (Supp Figure 1d). SIVgp120 protein and uncoupled BSA-blocked beads were included as negative controls for background subtraction. Plasma and saliva concentrations used in the array were chosen based on a dilution series (Supp Figure 2a). Briefly, antigen-coupled beads were incubated with the respective samples on a shaker overnight at 4°C, before being washed, and incubated with Phycoerythrin(PE)-conjugated detection antibodies (Southern Biotech) on a shaker for 2 hours at room temperature (RT). Beads were washed again and read on the Flexmap 3D.

Engagement of SARS-CoV-2 specific antibodies to Fc gamma receptors (FcγR) were measured using surrogate Fc gamma receptor dimers (FcγR2a, CD32; FcγR3a, CD16) as previously described (kind gift from Mark Hogarth and Bruce Wines) [16]. After incubation with samples, the washed beads were first incubated with surrogate FcγR dimers on a shaker for 2 hours at RT, washed again, and then incubated with Streptavidin-R-Phycoerythrin (SAPE; Thermo Fisher Scientific) on a shaker for a further 2 hours at RT. Finally, beads were washed and read on the Flexmap 3D.

### SARS-CoV-2 variant multiplex bead assay

To assess plasma (1:25600), saliva (1:12.5) and tear (1:25) antibody responses from booster and breakthrough cohorts, ancestral and variant (Alpha, Beta, Delta, Omicron BA.1, Omicron BA.2) whole ST and S1 were used to form a customized bead array (Supp Figure 2b). SARS-CoV-2 specific total IgG and IgA responses were assessed using biotin-conjugated detection antibodies (MabTech). As above, following incubation with samples, the washed beads were first incubated with the biotin-conjugated detection antibodies on a shaker for 2 hours at RT. Beads were then washed and incubated with SAPE for another 2 hours at RT, before being washed and read on the Flexmap 3D. The ability of SARS-CoV-2 variant specific plasma (1:12800) and saliva (1:12.5) antibodies to mediate FcγR engagements (FcγR2a, CD32; FcγR3a, CD16) were measured using the surrogate Fc-receptor dimers as described above.

### Variant RBD-ACE-2 inhibition bead assay

Neutralizing activity of plasma (1:800, 1:4000) and saliva (1:12.5) samples from booster and breakthrough cohorts against the SARs-CoV-2 variants of concern (Alpha, Beta, Delta, Omicron) were accessed using a surrogate RBD-ACE-2 inhibition assay. As previously described [1, 17], ancestral or variant RBD-coupled beads were incubated with avi-tagged biotinylated ACE2 in the presence of the respective plasma and saliva samples on a shaker for 2 hours at RT. Beads were washed and then incubated with SAPE on a shaker for 1 hours at RT. R-Phycoerythrin Biotin-XX conjugate (Thermo Fisher Scientific) was then added to the beads and incubated on a shaker for a further hour at RT. Finally, beads were washed and read on the Flexmap 3D. A nominal cutoff of 20% (depicted by dotted line) was also set as previously described [17]. Ancestral and variant specific RBD total IgG and IgA responses were also assessed using biotin-conjugated detection antibodies as described above (Supp Figure 2b).

### Analysis of viral RNA load by qPCR

For viral RNA extraction, briefly, 200 μL of sample was extracted with the QIAamp 96 Virus QIAcube HT kit (Qiagen, Germany) on the QIAcube HT System (Qiagen) according to manufacturer’s instructions. Purified nucleic acid was then immediately converted to cDNA by reverse transcription with random hexamers using the SensiFAST cDNA Synthesis Kit (Bioline Reagents, UK) as per manufacturer’s instructions. cDNA was used immediately in the rRT-PCR or stored at -20°C. Three microlitres of cDNA was added to a commercial real-time PCR master mix (PrecisionFast qPCR Master Mix; Primer Design, UK) in a 20 μL reaction mix containing primers and probe (final concentration of 0.9 mM primer and 0.2 mM probe, respectively). Samples were tested for the presence of SARS-CoV-2 RNA-dependent RNA polymerase (RdRp)/helicase (Hel), spike (S), and nucleocapsid (N) genes using previously described primers and probes [18, 19]. Thermal cycling and rRT-PCR analyses for all assays were performed on the ABI 7500 FAST real-time PCR system (Applied Biosystems, USA) with the following thermal cycling profile: 95C for 2 min, followed by 45 PCR cycles of 95C for 5 s and 60C for 25 s for N gene and 95C for 2 min, followed by 45 PCR cycles of 95C for 5 s and 55C for 25 s for RdRP/Helicase gene and S gene.

### Kinetics analysis

We used a piecewise model to estimate the activation time and growth rate of various immune responses (total IgG, IgA, and FcγR3aV) following breakthrough infections. The response variables had background levels subtracted by taking the mean of all the background values, and the threshold for detection was set at two standard deviations above the background responses. The model of the immune response *y* for subject *i* at time *y*_*i*_ can be written as:

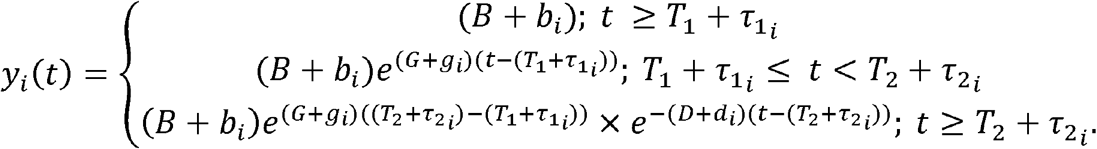

The model has 5 parameters: baseline level (B), growth rate (G), timing on onset of growth (T_1_), decay (D), and time of peak (T_2_). For a period before 1, we assumed a constant baseline value for the immune response (which is higher than or at the background level). After the activation time *T*_1_, the immune response will grow at a rate Of *G* until *T*_2_. From *T*_2_, the immune response will decay at a rate of *D*. For each subject *i*, the parameters were taken from a normal distribution, with each parameter having its own mean (fixed effect). A diagonal random effect structure was used, where we assumed there was no correlation within the random effects. The model was fitted to the log-transformed data values, with a constant error model distributed around zero with a standard deviation *σ*. To account for the values less than the limit of detection, a censored mixed effect regression was used to fit the model. Model fitting was performed using MonolixR2019b.

### Statistical analysis

Statistical analysis was performed with GraphPad Prism 9 (GraphPad Software). To transform the data in percentages for use in the radar plots (Figure 1b; Supp Figure 3a), the median of each cohort/timepoint’s antigen-specific detector-specific MFI was divided by the antigen-specific MFI from the 98^th^ percentile for that detector (98^th^ percentile was chosen to minimize the impact of outliers on the data transformation). Antibody levels between cohort/timepoints were compared using Mann-Whitney tests or Friedman tests, with corrections for multiple comparisons as required. Spearman’s rank correlation analyses were performed to study associations between antibody signatures.

**Figure 1:**
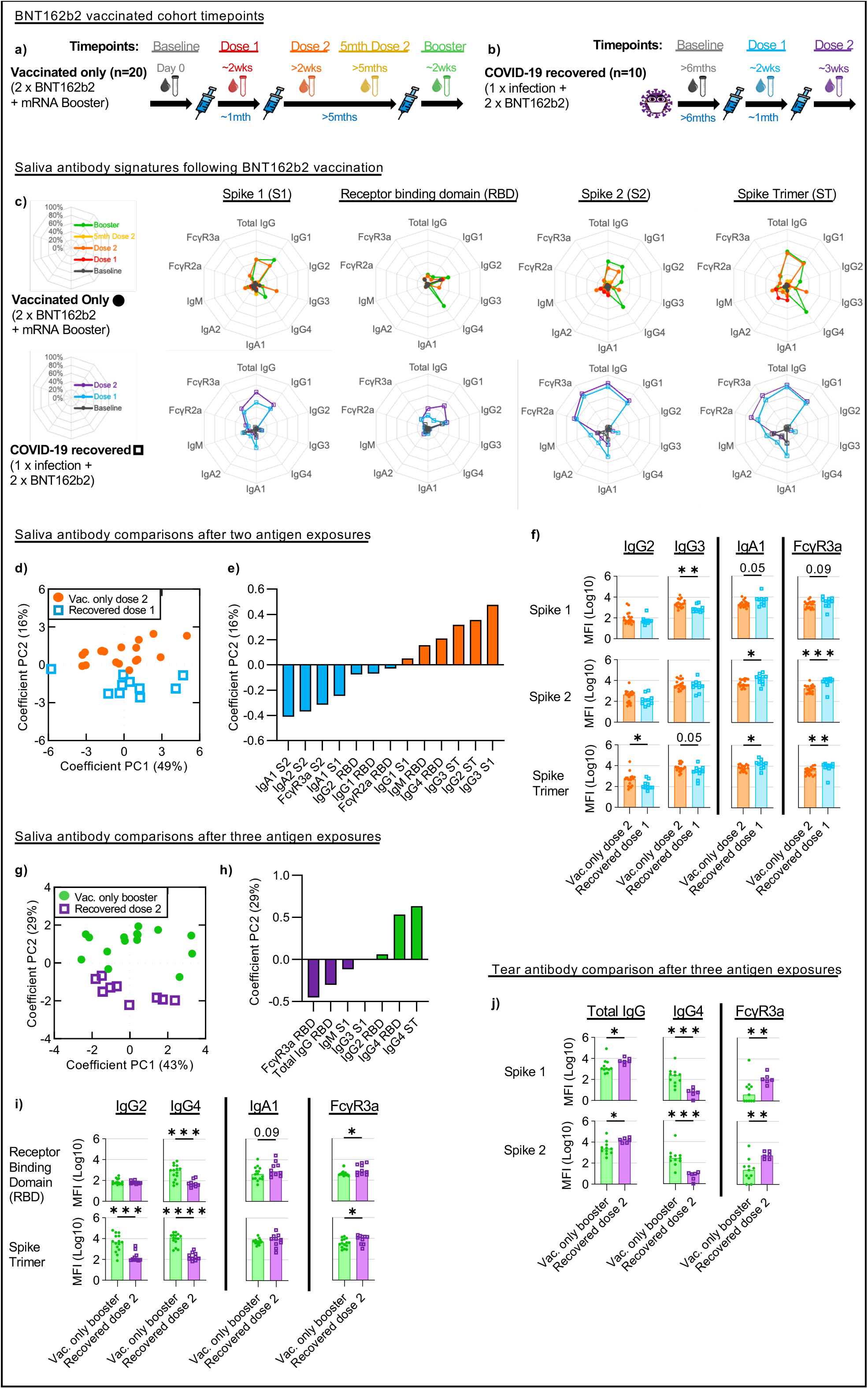
Salivary and tear antibodies from COVID-19 recovered individuals show stronger IgA and Fc_γ_R engagement responses after COVID-19 mRNA vaccination. Paired saliva and plasma samples were collected pre- and post-mRNA vaccination from vaccinated only **(a)** and COVID-19 recovered **(b)** individuals at the indicated time-points. Saliva antibody isotype and subclass responses from both cohorts against the various SARS-CoV-2 spike antigens were compiled into respective radar plots **(c)**. The individual median antibody isotype/subclass response for each spike antigen was transformed into percentages using the antigen-specific MFI from the 98^th^ percentile for that detector (98^th^ percentile was chosen to minimize the impact of outliers on the data transformation). PCA of all 40 antibody features for vaccinated only (closed circles) and COVID-19 recovered (open squares) individuals after two **(d)** and three **(g)** antigen exposures. Loading plots and bar graphs describe the key differences between both cohorts after two **(e**,**f)** and three **(h**,**i)** antigens exposure. Major tear antibody features after three antigen exposures are also illustrated in bar graphs **(j)**. Statistical significance was calculated using the two-tailed Mann-Whitney *U* test and where significant or trending significance, *p*-values were reported (* p≤0.05; ** p≤0.01; *** p≤0.001; **** p≤0.0001).

### Data normalization for Multivariable multiplex analysis

Positive control antigens (Influenza A H1N1 hemagglutinin) were removed from the database prior to analysis. Negative control antigens (SIV gp120, BSA) were background subtracted for each individual antigen-detector pair. If the any feature contained any negative values, the entire dataset was right shifted by adding the minimum value for that feature back to all samples within that feature. Right-shifted data were log-transformed using the following equation, where x is the right-shifted data and y is the right-shifted log-transformed data: y = log10(x + 1) to achieve normal distribution. Data were furthered normalized by mean centering and variance scaling each feature using the z-score function in MATLAB in the subsequent multivariate analyses as previously described [16].

### Multivariable methods for identification of key antibody features

A Least Absolute Shrinkage and Selection Operator (LASSO) penalized logistic regression model was used to identify the minimal set of features that differentiated between COVID-19 recovered and COVID-19 vaccinated only vaccinees [20]. The feature selection stability was defined as the proportion of times that a feature was selected when the model was repeatedly fitted to 1000 resampled subsets of data as previously described[21]. Principal Component analysis (PCA) was then performed. Two-dimensional score plots were generated to visually assess separation between groups. All analysis was conducted using the Statistics and Machine Learning Tool on MATLAB, data were extracted and figures were graphed using GraphPad Prism.

## Results

### Mucosal IgG4 is elevated after third mRNA vaccine dose

Individuals infected with COVID-19 prior to SARS-CoV-2 vaccinations (COVID-19 recovered vaccinees) elicit stronger systemic total IgG and neutralization responses, than those induced only by SARS-CoV-2 vaccination alone [2, 4-6]. Furthermore, prior mucosal exposure due to SARS-CoV-2 infection primes improved mucosal total IgG and IgA responses resulting from subsequent IM COVID-19 vaccinations [2, 4-6]. However, few studies have delved into the antibody subclass expression, as well as antibody-mediated functional responses, particularly at the mucosa, within such vaccinees.

To address this, we profiled SARS-CoV-2-specific salivary antibody isotypes, subclasses and functional responses from both COVID-19 vaccinated only vaccinees receiving up to 3 mRNA vaccines (vaccinated only; 2 x BNT162b2 + 1 x mRNA booster), and COVID-19 recovered vaccinees receiving up to 2 mRNA vaccines (COVID-19 recovered; 1 x prior COVID infection + 2 x BNT162b2) (Figure 1a-c). The multiplex array used contained both ancestral SARS-CoV-2 receptor binding domain (RBD) and Spike 1 (S1) to study novel responses made against SAR-CoV-2, as well as ancestral SARS-CoV2 Spike 2 (S2) and whole Spike trimer (ST) which detect cross-reactive responses conserved across other coronaviruses (Supp Figure 1d) [22].

After two antigen exposures, we detected marked differences in salivary antibody signatures between COVID-19 recovered (1 x prior infection + 1 x BNT162b2) and vaccinated only cohorts (2 x BNT162b2) (Figure 1d, e). As compared to the vaccinated only cohort, COVID-19 recovered vaccinees developed better salivary IgA responses (Figure 1e, f; Supp Figure 3a). However, we noted that these salivary IgA responses were biased towards the more conserved Spike proteins S2 and ST responses (p≤0.05) instead of the novel S1 or RBD (Figure 1e, f; Supp Figure 3a). Importantly, salivary antibodies from COVID-19 recovered vaccinees displayed higher FcγR engagement following two antigen exposures (1 x prior infection + 1 x BNT162b2) as compared to the vaccinated only cohort (2 x BNT162b2), also primarily against the conserved antigens S2 and ST (p≤0.01) (Figure 1e, f; Supp Figure 3a). On the other hand, salivary IgG subclass responses in vaccinated only vaccinees (2 x BNT162b2) aligned with that observed in plasma, with subtle increases across both IgG2 against the conserved ST (p≤0.05), as well as IgG3 responses against the novel S1 (p≤0.01) respectively (Figure 1e, f; Supp Figure 3a, 4a-d).

Diverse differences in salivary antibodies were also detected following the third antigen exposures between COVID-19 recovered (1 x prior infection + 2 x BNT162b2) and vaccinated only (2 x BNT162b2 + 1 x mRNA booster) cohorts (Figure 1g, h). In contrast to their first mRNA vaccination, there was a decline in salivary IgA response among the COVID-19 recovered vaccinees after their second mRNA vaccination (Figure 1c, i; Supp Figure 3b, d). This supports previous observations that salivary IgA responses dip after the second IM vaccine dose and highlights a potential need for repeated mucosal antigen exposures to induce or maintain robust local IgA responses [6]. Conversely, salivary total IgG levels remained higher in the COVID-19 recovered cohort (1 x prior infection + 2 x BNT162b2) than the vaccinated only cohort (2 x BNT162b2 + 1 x mRNA booster) after three antigen exposures, largely driven by IgG1 responses (Figure 1c, h; Supp Figure 3b). More importantly, COVID-19 recovered vaccinees still induced better antibody-mediated Fc-engagement than vaccinated only vaccinees across multiple spike antigens (p≤0.05) (Figure 1c, h, i; Supp Figure 3b).

In contrast, salivary IgG subclass responses in vaccinated only vaccinees (2 x BNT162b2 + 1 x mRNA booster) once again mimicked that observed in plasma after three antigen exposures, with strongly elevated IgG2 (p≤0.05) and IgG4 (p≤0.001) responses detected across multiple SARS-CoV-2 spike antigens, particularly the more conserved S2 and ST (Figure 1c, h, i; Supp Figure 3b, 4a, e-g). Rises in IgG2 and particularly IgG4 responses in blood post-mRNA vaccination have been previously described following repeated mRNA vaccinations and were instead absent following repeated vaccinations with adenoviral vectors [23, 24]. As such, here we also tested the salivary responses in vaccinated only vaccinees who had received 2 adenoviral vector vaccines prior to their mRNA booster (2 x ChAdOx1 nCoV-19 + 1 x mRNA booster; total 3 x antigen exposures) (Supp Figure 1a, 5a). Unsurprisingly, while the mRNA booster enhanced total IgG levels in saliva, it did not induce a detectable IgG4 responses, mimicking that observed in plasma (Supp Figure 5b, c).

IgG subclass switching to IgG2 and subsequently IgG4 is often thought to be a compensatory mechanism against over-inflammation due to their relatively poor ability to engage Fc Receptors [25]. Irrgang et al. recently demonstrated that the rise in IgG4 antibodies after repeated COVID-19 vaccinations coincided with a decrease in SARS-CoV-2 specific antibody-mediated functional responses in blood [24]. Here, while we did detect negative correlations between both IgG2/IgG4 responses against Fcγ2aR/Fcγ3aR engagements in plasma from vaccinated only vaccinees after their mRNA boosters (2 x BNT162b2 + 1 x mRNA booster), this pattern was not replicated in paired saliva samples (Supp Figure 3f, g; 4h, i). This suggests that the presence of elevated IgG2 and IgG4 in saliva might not be sufficient to dampen antibody-mediated functional responses at the mucosa in vaccinees receiving multiple mRNA vaccines.

To corroborate the mucosal antibody signatures observed in saliva following vaccination, we further explored if similar antibody signature would also be detected in tear fluid. Despite being found in different mucosal sites, both tear fluid and saliva serve as the first line of defense against aerosolized SARS-CoV-2. Similar to salivary antibody responses, only three doses of mRNA vaccines induced elevated anti-spike IgG4 levels in tear fluid (Figure 1j; Supp Figure 3c). Vaccinees with prior COVID-19 infection also had higher levels of total IgG and FcγR3a responses in tear fluid after three antigen exposures (1 x prior infection + 2 x BNT162b2) (Figure 1j; Supp Figure 3c). Additionally, mucosal IgA responses in tear fluid of COVID-19 recovered vaccinees peaked after the first vaccine dose and waned after the second dose (Supp Figure 3e). These findings suggest that mucosal antibody responses to IM vaccination are conserved across different mucosal sites.

### Prior COVID-19 infection induces weak salivary neutralization of RBD-ACE2 interactions

Neutralizing activity is key in both protecting from SARS-CoV-2 infection and preventing severe disease [26]. Mucosal neutralizing antibodies have been shown to protect against viral challenge and are the goal of COVID-19 mucosal vaccines currently in development [27]. Salivary neutralizing activity can be detected in individuals with hybrid immunity, although it may be more limited than responses in plasma and more technically challenging to measure [28]. Here, we compared the ability of plasma and salivary antibodies from our vaccinated only and COVID-19 recovered cohorts to inhibit ACE2 binding to a series of RBDs, including ancestral SARS-CoV2 and the various variants of concern (VoC). This RBD-ACE2 surrogate assay correlates well with cell-based live virus microneutralization assay, while avoiding cell-based complications arising from the use of non-sterile saliva, making it suitable for interrogating our saliva samples [1, 17].

Following a mRNA booster, plasma from vaccinated only vaccinees (2 x BNT162b2 + 1 x mRNA booster) strongly inhibited ACE-2 binding to both ancestral and pre-Omicron VoC RBDs (p≤0.0001) (Figure 2a). Inhibition of ACE2 binding to Omicron BA.1 and BA.2 RBDs were much more modest (ο BA.1: not significant; ο BA.2: p≤0.01 respectively). Similarly, plasma from COVID-19 recovered vaccinees after their first mRNA vaccine showed robust inhibition of ACE-2 binding to RBDs from both ancestral and pre-Omicron VoCs (p≤0.01) (Figure 2b) However, consistent with that observed with the vaccinated only cohort, Omicron BA.1 and BA.2 RBD-ACE2 inhibition responses in plasma were weaker (ο BA.1: not significant; ο BA.2: p≤0.05 respectively), with minimal improvement noted even after the second mRNA vaccine (ο BA.1: not significant; ο BA.2: p≤0.01 respectively) (Figure 2c).

**Figure 2:**
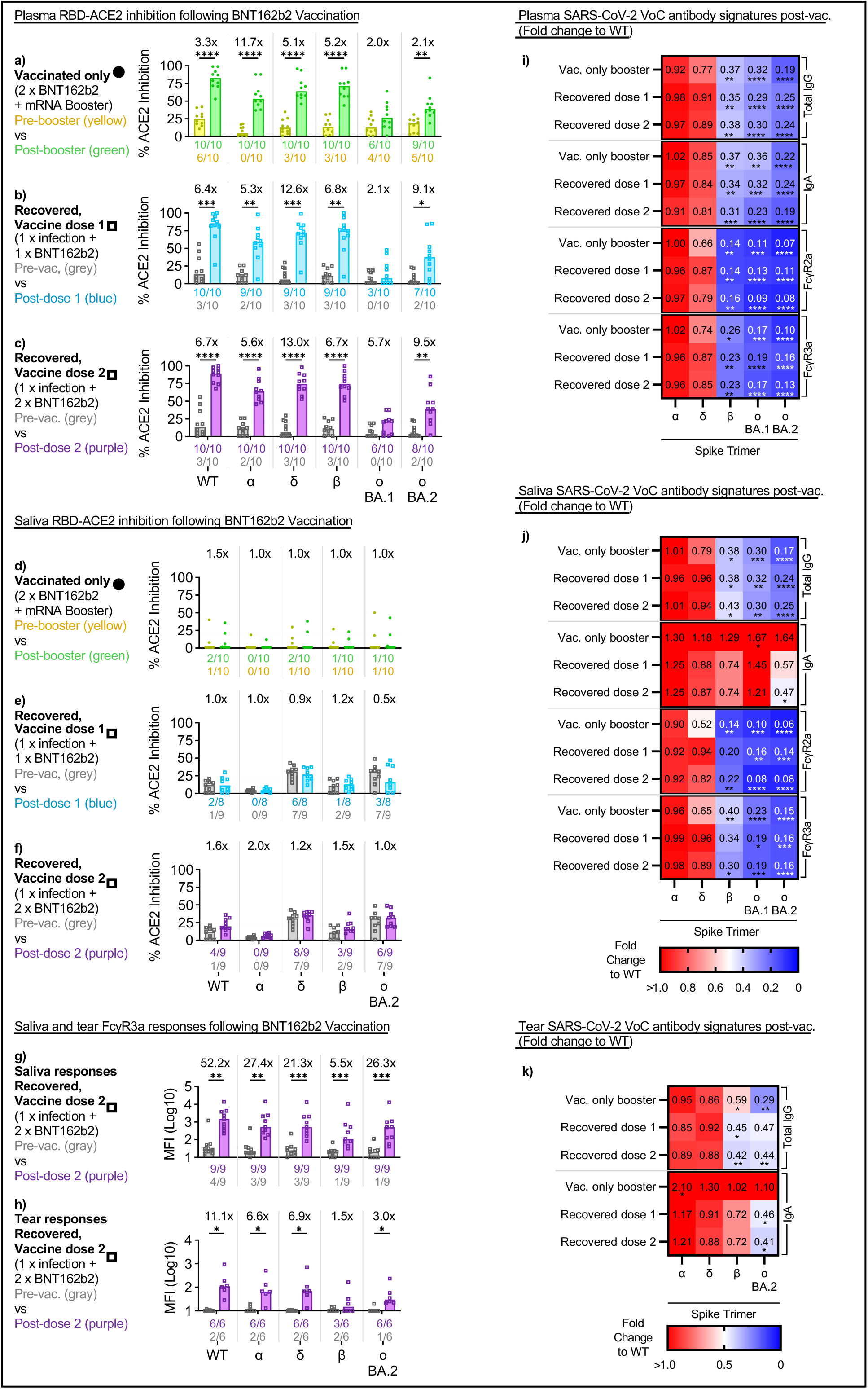
Ancestral imprinting limits cross-reactive responses against VoCs elicited by COVID-19 mRNA vaccination. Bar graphs depict the plasma **(a-c)** and salivary **(d-f)** inhibition of RBD-ACE2 interactions against the ancestral wildtype (WT) SARS-CoV-2 or the VoCs (α, Alpha; δ, Delta; β, Beta; ο BA.1, Omicron BA.1; ο BA.2, Omicron BA.2) by vaccinated only **(a, d)** and COVID-19 recovered individuals **(b, c, e, f)** respectively. Bar graphs also display the salivary **(g)** and tear **(h)** Fcγ3a responses against WT SARS-CoV-2 or the VoCs in COVID-19 recovered individuals following two doses of mRNA vaccines. Fold changes listed above the bar graphs were calculated for post-booster (green) and post-vaccination responses (blue, purple) over their respective pre-booster (yellow) and pre-vaccination responses (grey) for each cohort and antigen. The number of individuals with detectable responses above the assay threshold (dotted line) (RBD-ACE2: arbitrary 20%; Fcγ3a: pre-pandemic average) at either timepoint were listed under the bar graphs in their respective colours. Significant differences between both timepoints were calculated using the two-tailed Mann-Whitney *U* test, followed by Bonferroni-Dunn’s test for multiple comparisons. Heat map illustrate the VoC-specific Spike Trimer plasma **(i)** salivary **(j)** and tear **(k)** antibody responses post-booster and post-vaccination (dose 1 or 2) for the vaccinated only and COVID-19 recovered cohorts respectively. The median antibody response for each VoC spike was described as a fold change to the wildtype spike. Statistical significance was calculated using Friedman’s test followed by Dunn’s test for multiple comparisons. Where significant, *p*-values were reported (* p≤0.05; ** p≤0.01; *** p≤0.001; **** p≤0.0001).

Expectedly, we did not find meaningful RBD-ACE2 inhibitory activity in saliva from vaccinated only vaccinees even after their mRNA booster (2 x BNT162b2 + 1 x mRNA booster) (1-2% median inhibition across WT and VoCs), despite detectable total IgG and IgA antibodies against RBD (Figure 1c, i; Supp Figure 3b) [1]. In contrast, while we detect weak RBD-ACE2 inhibitory activity in the saliva from COVID-19 recovered vaccinees even prior to vaccination, these responses did not improve significantly, even after their second mRNA vaccine dose (1 x prior infection + 2 x BNT162b2) (Figure 2e, f). These findings support the notion that salivary neutralizing antibodies are induced following local antigen exposure at the mucosa but not by IM mRNA vaccination alone.

### Broad cross-reactivity in mucosal IgA against VoC spike trimers

Despite the low levels of mucosal neutralizing antibodies, we have shown above that antibodies mediating FcγR engagement were detectable in saliva and tear fluid, particularly in COVID-19 recovered vaccinees after their second mRNA vaccine (1 x prior infection + 2 x BNT162b2) (Figure 1c, j; Supp Figure 3a-c). These FcγR responses in saliva and tear fluid were enhanced in COVID-19 recovered vaccinees after their second mRNA vaccination and could target a range of VoC spikes (Figure 2g, h). However, it was also noticeable that despite similar levels of pre-vaccination responses, the largest gains after ancestral vaccination were unsurprisingly ancestral-centric mucosal humoral responses (Figure 2g, h).

SARS-CoV-2 antibody breadth has been shown to passively increase one year following SARS-CoV-2 infection as a result of continued evolution of anti-SARS-CoV-2 antibodies targeting the viral spike [29]. However, repeatedly exposing COVID-19 recovered vaccinees to the ancestral antigen through vaccination could bias ancestral-centric responses and diminish efforts towards developing broader antibody responses capable of recognizing newer VOCs. To explore this, we compared the systemic and mucosal humoral responses made by COVID-19 recovered vaccinees after their first and second mRNA vaccine doses to that from vaccinated only vaccinees. Antibody responses to the respective VoC spikes trimer were quantified and their relative abundance were compared against the ancestral wildtype (WT) spike (fold change over WT).

Broadly, the abundance in cross-reactive antibody responses against the various VoCs followed their hierarchy of escape mutations. Cross-reactive total IgG and FcγR responses against the much conserved Alpha spike trimers were mostly comparable with the ancestral WT spike trimer in both plasma and saliva across all cohorts (Figure 2i, j). On the other hand, cross reactive responses to Delta spike trimer were less abundant in booster vaccinated only vaccinees in comparison to COVID-19 recovered vaccinees, especially for Fcγ2aR and Fcγ3aR responses for both plasma and saliva (Figure 2i, j).

However, even more strikingly, the relative abundance of cross-reactive total IgG in all cohorts against the less conserved Omicron (BA.1, BA.2) (blue on heat map) remained smaller than the more conserved Alpha and Delta VoCs (red on heat map) in both plasma (ο BA.1: 0.29 – 0.32; ο BA.2: 0.19 – 0.25; p≤0.0001) and saliva respectively (ο BA.1: 0.30 – 0.32; ο BA.2: 0.17 – 0.25; p≤0.01) after vaccination (Figure 2i, j). The relative spread of cross-reactive plasma and salivary FcγR responses, as well as plasma IgA against the Omicron also largely followed a similar trend across all cohorts following vaccination (Figure 2i, j). As such, while vaccination with the ancestral WT spike did increase SARS-CoV-2-specific antibodies, these were largely biased towards ancestral-centric responses.

To the contrary, salivary IgA from COVID-19 recovered vaccinees displayed a much broader range of cross-reactivity against Beta and Omicron (BA.1, BA.2) spike trimers after their first and second mRNA vaccines. This trend of larger abundance in cross-reactive mucosal IgA capable of targeting Omicron was also corroborated in our analysis with tear fluid from COVID-19 recovered vaccinees following their first and second mRNA vaccines (Figure 2k). As such, the stimulation of broadly cross-reactive mucosal IgA could be key in establishing a robust protective barrier against SARS-CoV-2 infections at the mucosa by the newly emerging VoCs.

### Salivary IgA and FcγR3a antibody-mediated responses are enhanced during breakthrough infections

Given that prior COVID-19 infection could influence both systemic and more importantly, mucosal humoral responses following vaccination, we next wanted to explore how prior vaccination impacted systemic and mucosal humoral responses during COVID-19 breakthrough infections. To address this, we expanded our studies to breakthrough COVID-19 cohorts and collected serial saliva and plasma samples from vaccinated individuals with acute COVID-19 from two different VoC waves (Delta, Omicron BA.2) in Victoria, Australia (Figure 3a, Supp Figure 1b, c) [8, 13, 14]. SARS-CoV-2 viral load (nasal swabs) from both VoC waves had comparable viral loads as previously reported (Supp Figure 6a-b), which titered out by two weeks [8]. As such we compared antibody responses up to two weeks, to determine factors that could be associated with viral clearance. Surprisingly, despite having high detectable viral loads, only a minority of individuals with breakthrough infections developed neutralizing antibodies in their saliva (Figure 3b, c; Supp Figure 6c). Most individuals with breakthrough infections failed to produce a response above our 20% assay cutoff, regardless of breakthrough wave (Delta, Omicron BA.2) (Figure 3b, c; Supp Figure 6c).

**Figure 3:**
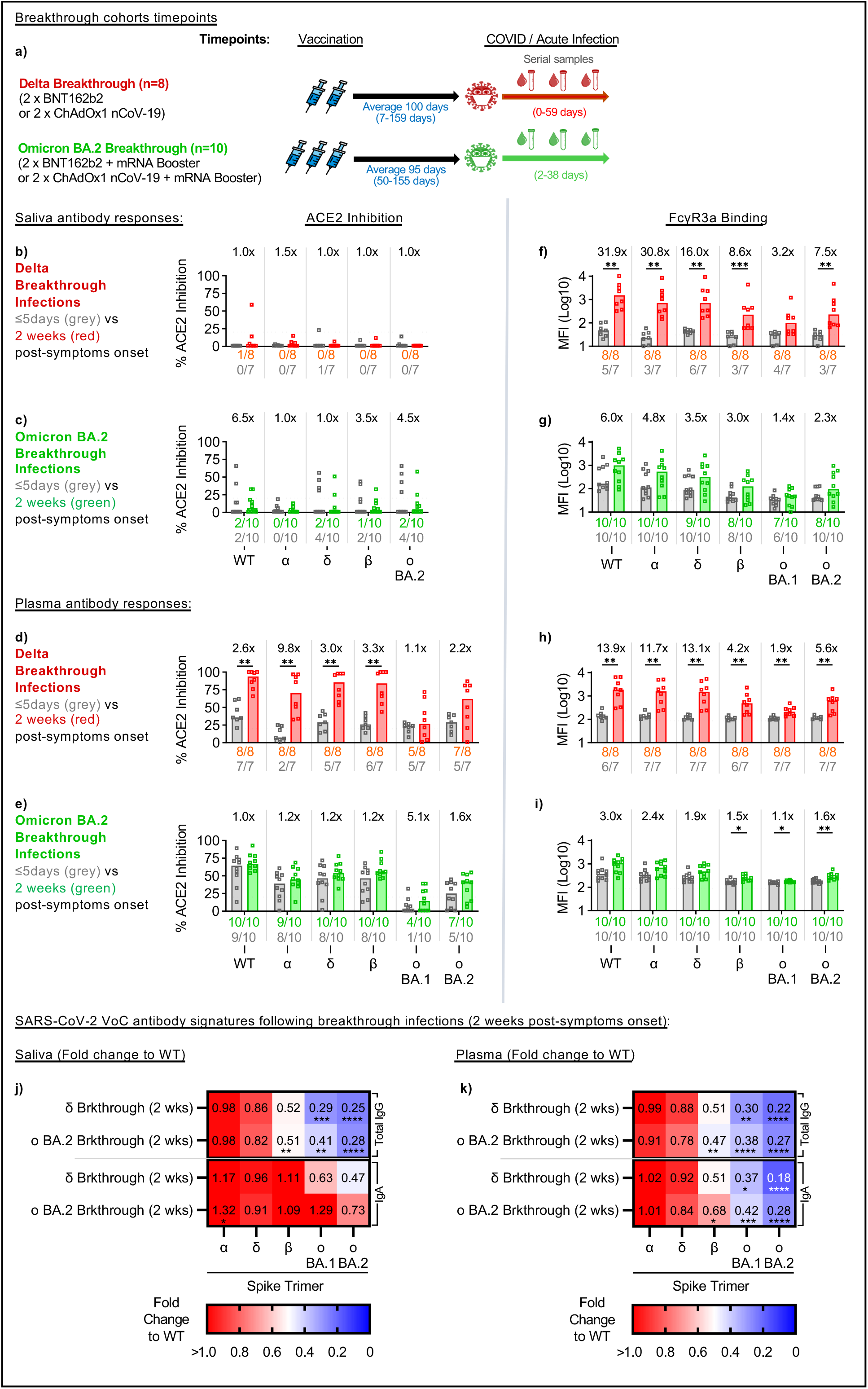
Pre-existing vaccine-induced immunity modulates variant-specific antibody responses during breakthrough infections. Paired saliva and plasma samples were serially collected from individuals over the course of their Delta or Omicron BA.2 breakthrough infections **(a)**. Bar graphs depict the salivary **(b, c)** and plasma **(d, e)** inhibition of RBD-ACE2 interactions against the ancestral wildtype (WT) SARS-CoV-2 or the VoCs (α, Alpha; δ, Delta; β, Beta; ο BA.1, Omicron BA.1; ο BA.2, Omicron BA.2) by individuals with Delta **(b, d)** and Omicron BA.2 breakthrough **(c, e)** infections respectively. Similarly, bar graphs depict the engagement of FcγR3a by salivary **(f, g)** and plasma **(h, i)** antibodies by individuals with Delta **(f, h)** and Omicron BA.2 breakthrough **(g, i)** infections respectively. Fold changes listed above the bar graphs were calculated for responses 2 weeks post-symptom onset (Delta: red; Omicron BA.2: green) over respective responses earlier during infection (grey; ≤5 days post-symptom onset) for each cohort and antigen. The number of individuals with detectable responses above the assay threshold (dotted line) (RBD-ACE2: arbitrary 20%; FcγR3a: pre-pandemic average) at either timepoint were listed under the bar graphs in their respective colours. Significant differences between both timepoints were calculated using the two-tailed Mann-Whitney *U* test followed by Bonferroni-Dunn’s test for multiple comparisons. Heat map illustrate the VoC-specific Spike Trimer salivary **(j)** and plasma **(k)** antibody responses for Delta and Omicron BA.2 breakthrough cohorts 2 weeks post symptom onset. for the vaccinated only and COVID-19 recovered cohorts respectively. The median antibody response for each VoC spike was described as a fold change to the wildtype spike. Statistical significance was calculated using Friedman’s test followed by Dunn’s test for multiple comparisons. Where significant, *p*-values were reported (* p≤0.05; ** p≤0.01; *** p≤0.001; **** p≤0.0001).

To study FcγR engagement, we focused on FcγR3a responses as we had previously noted that they were stronger and less impacted by ancestral imprinting than FcγR2a (Figure 2i, j) In contrast to neutralizing antibodies, most individuals with Delta or Omicron BA.2 COVID-19 breakthrough infections had detectable FcγR engagement responses in saliva (Figure 3f, g; Supp Figure 6d). Individuals with Delta breakthrough infections had two doses of COVID-19 vaccines and displayed low levels of cross-reactive salivary antibodies against the various VoC spikes during early infection, with only about half of the cohort recording responses above pre-pandemic controls (dotted line) (Figure 3f; Supp Figure 6d). However, after two weeks, robust generation of salivary antibody-mediated functional responses were detected against the Delta variant spike (16-fold increase, p≤0.01) (Figure 3f; Supp Figure 6d). The effects of ancestral imprinting were also noticeable, with the ancestral WT (31.9-fold increase, p≤0.01) and more conserved Alpha variant spikes (30.8-fold increase, p≤0.01) gaining the largest increases in FcγR responses two weeks post-symptom onset (Figure 3f; Supp Figure 6d).

In contrast, all individuals with Omicron BA.2 breakthroughs had their mRNA boosters (3 x vaccine doses) and displayed higher levels of pre-existing salivary antibodies capable of engaging FcγR early in infection as compared to the Delta breakthrough cohort (Figure 3g; Supp Figure 6d). Smaller fold-increases were observed for Omicron BA.2 infections two weeks post-symptom (ο BA.2: 2.3-fold increase), with the largest differences still being ancestral-centric responses (WT: 6-fold increase) (Figure 3g; Supp Figure 6d). The Omicron BA.2 breakthrough cohort also achieved an overall lower maximal FcγR response across all variants tested as compared to that observed with Delta breakthrough infections Figure 3f, g; Supp Figure 6d).

Furthermore, while the relative abundance of salivary total IgG responses remained largely ancestral-centric, there was a wider spread of cross-reactive salivary IgA responses that were elicited in both breakthrough cohorts after two weeks of symptom (Figure 3j). This supports our observation that while levels of total IgG were associated with viral clearance in the systemic compartment (plasma) (Supp Figure 6g, h), IgA levels correlated better with viral clearance in the mucosal compartment (saliva), particularly with Omicron BA.2 breakthrough infections (Supp Figure 6i, j).

Taken together, our mucosal data suggest that despite limited neutralizing activity against novel SARS CoV-2 antigens, salivary antibodies targeting FcγR engagement, as well as salivary IgA, could still play key roles for localized cross-reactive protection at the mucosa. However, ancestral-centric pre-existing immunity may also influence the type and magnitude of salivary FcγR engagement antibody responses made during breakthrough infections.

### Pre-existing vaccine immunity modulates systemic neutralizing and functional antibody responses during breakthrough infections

In comparison, systemic antibody responses (plasma) displayed better neutralizing activity. Individuals with Delta breakthroughs (2 x COVID-19 vaccines) started out with lower levels of cross-reactive plasma antibodies against the Delta variant RBD (29% median RBD-ACE2 inhibition) early in infection (Figure 3d; Supp Figure 6e). After two weeks post-symptom onset, the robust generation of Delta-specific antibodies led to a 3-fold increase in RBD-ACE2 inhibition (Figure 3d; Supp Figure 6e). Notably, significant increases in ACE2 inhibition were also observed for the ancestral strain as well as more conserved pre-Omicron variants (Alpha, Delta, Beta) (p≤0.01). In contrast, little difference to ACE2 inhibition was seen with the more immune-escaped Omicron BA.1 RBD (1.1-fold increase in RBD-ACE2 inhibition).

Individuals with Omicron BA.2 breakthroughs all had their mRNA boosters (3 x vaccine doses) but were still infected despite having higher levels of pre-existing ancestral-centric SARS-CoV-2 antibodies (65% median RBD-ACE2 inhibition against WT), highlighting the immune evasiveness of the variant (Figure 3e; Supp Figure 6e). Smaller increases in RBD-ACE2 inhibition (1 to 1.2-fold increases) particularly against the ancestral wildtype and the more conserved pre-Omicron variants (Alpha, Delta, Beta) were observed after two weeks, despite having a lower maximal response than that from the Delta wave (67% vs 94% median RBD-ACE2 inhibition against WT) (Figure 3e; Supp Figure 6e). In contrast, the largest growth in ACE2-inhibition after two weeks were against the Omicron variants (ο BA.1: 5.1-fold increase; ο BA.2: 1.6-fold increase), although they were still lower than cross-reactive responses induced from the Delta breakthroughs (15% vs 27% and 41% vs 62% median RBD-ACE2 inhibition against ο BA.1 and ο BA.2 respectively)(Figure 3e; Supp Figure 6e).

Systemic FcγR engagement responses trended similar to that described above for the mucosa. During early Delta breakthrough infection, most individuals had minimal levels of antibodies capable of FcγR3a engagement, like that of pre-pandemic controls (dotted line) (Figure 3h; Supp Figure 6f). However, after two weeks, significant increases to FcγR engagements were noticed across all VoCs, including Omicron (p≤0.01). The largest increase observed were ancestral-centric responses with the ancestral wildtype (13.9-fold increase), as well as the more conserved Alpha (11.7-fold increase) and Delta spikes (13.1-fold increase).

On the other hand, pre-existing levels of FcγR engagement from samples collected early in Omicron BA.2 infections remained above that found in uninfected pre-vaccination controls (Figure 3i; Supp Figure 6f). Smaller ancestral-centric fold-increases were observed for BA.2 infections 2 weeks post-symptom onset, achieving an overall lower maximal response across all variants tested as compared to that observed with Delta breakthrough infections (Figure 3e; Supp Figure 6f). Notably, the growth in plasma FcγR responses against the range of VoCs by 2 weeks post-symptom onset were also mostly smaller than those changes elicited at the mucosa (Figure 3f-I; Supp Figure 6d, f).

Taken together, our data support the notion that higher levels of pre-existing plasma antibodies could negatively influence the magnitude of systemic humoral responses elicited during breakthrough infections despite comparable viral loads as measured through nasal swabs. The effects of imprinting from the ancestral strain also appear to be more obvious with FcγR engagement responses, possibly due to the larger involvement of conserved cross-reactive antibodies, as compared to neutralization.

### Rapid recall of salivary antibodies in Omicron Breakthrough compared to plasma

Since pre-existing SARS-CoV-2 immunity could modulate the magnitude of antibody responses detected two weeks after the onset of breakthrough infections, we next investigated if variations in antibody kinetics could explain the differential rise observed in systemic and mucosal antibodies targeting the respective spike antigens. Here, we modelled the dynamics of antibody features (total IgG, IgA and Fcγ3aR) targeting the ancestral WT, Delta, Omicron BA.1 and Omicron BA.2 Spike 1 (S1) and Spike Trimer (ST) using serial plasma and saliva samples collected from both respective COVID-19 breakthrough cohorts for up to 40 days.

Regardless of VoC waves, antibody responses towards the ancestral WT spike (black line) largely dominated in both plasma and saliva even up to 40 days (Figure 4a, c; Supp Figure 7a, c). The magnitude of responses to Delta (red line), which is more similar to the ancestral WT, was also usually higher than those for Omicron (BA.1 and BA.2; blue and green lines respectively) even for the Omicron breakthrough cohorts. These observations corroborate with our above findings that antibody responses made during acute breakthrough infection could be ancestral-centric and largely driven by cross-reactive responses instead.

**Figure 4:**
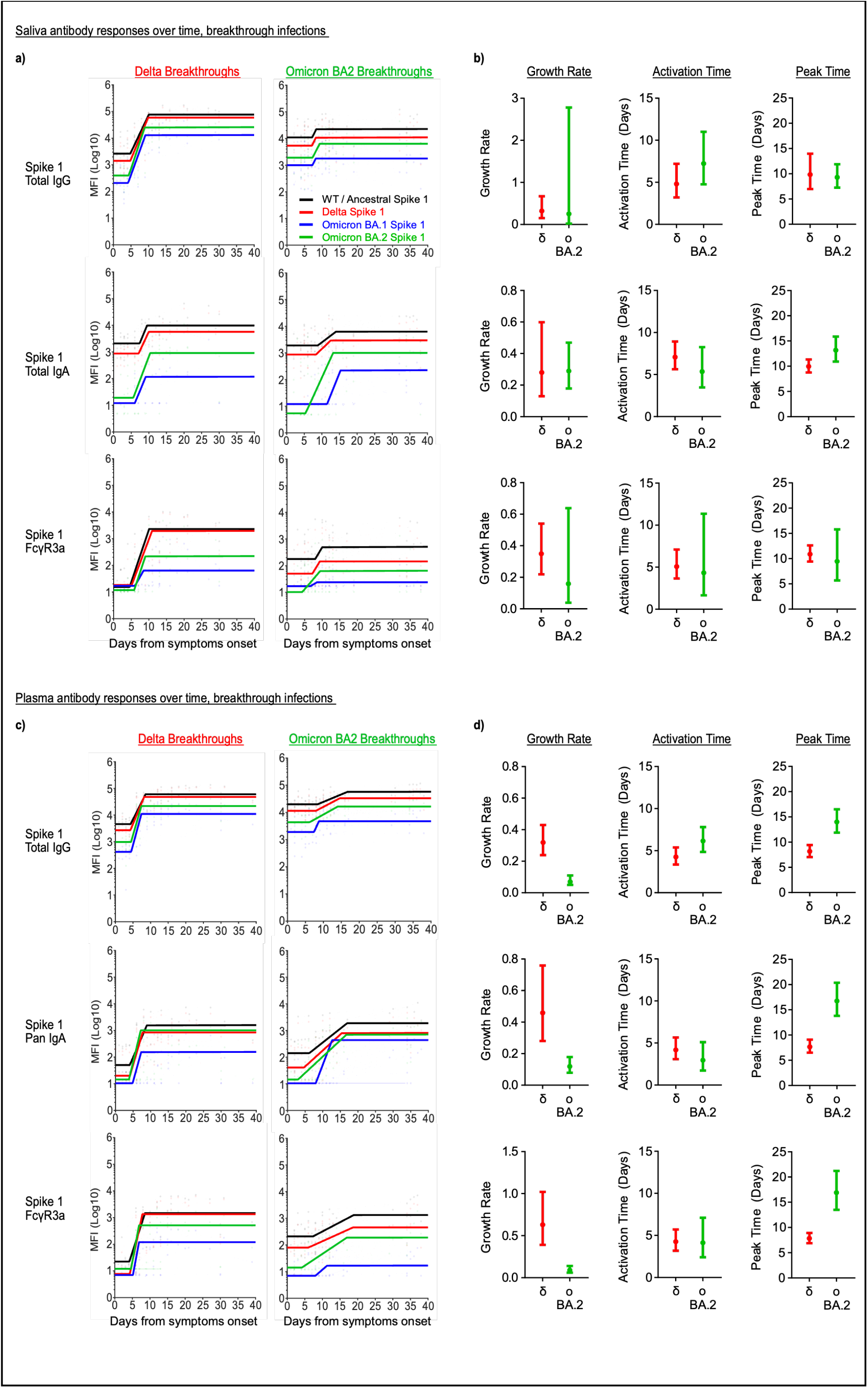
Plasma but not salivary antibody responses displayed delayed kinetics during Omicron BA.2 breakthrough infection. Modelled kinetic curves (WT: black; Delta: red; Omicron BA.1: blue; Omicron BA.2: green) describe the ancestral wildtype and variant-specific Spike 1 antibody responses from serially collected saliva **(a)** and plasma **(c)** samples during Delta or Omicron BA.2 breakthrough infection for up to 40 days post-symptom onset. Connected dotted lines indicate serial samples from the same individual. Lines with open circles at the bottom of each graph reflect samples that were excluded from the model for being below the threshold of detection (2 S.D. background readings). Dot plots displaying 95% confidence intervals beside each row of kinetic curves list the calculated growth rate, time to activation and time to peak of variant-specific salivary **(b)** and plasma **(d)** responses (Delta: red; Omicron BA.2: green) by their respective breakthrough cohorts (eg: Delta variant responses during the Delta breakthroughs).

Despite larger variations particularly with the Omicron BA.2 responses, the growth rate, time of activation, and time of peak for salivary total IgG, IgA and Fcγ3aR responses against the respective breakthrough variant’s S1 and ST were comparable between the Delta and Omicron BA.2 breakthrough cohorts (with overlapping 95% confidence intervals) (Figure 4b; Supp Figure 7b). The time of activation for plasma IgG, IgA and Fcγ3aR responses against the respective breakthrough variant’s S1 were also comparable between the Delta and Omicron BA.2 breakthrough cohorts (with overlapping 95% confidence intervals) (Figure 4d). However, a delay in time of activation was detected in plasma IgA and Fcγ3aR responses against Omicron BA.2 ST by the Omicron BA.2 breakthrough cohort, as compared to Delta responses in the Delta breakthrough cohort (no overlapping 95% confidence intervals) (Supp Figure 7d).

In contrast, the growth rate for total IgG, IgA and Fcγ3aR responses in plasma against Omicron BA.2 S1 and ST in the Omicron BA.2 breakthrough cohort (ο BA.2 S1: 0.07, 0.12, 0.09 respectively) was much poorer than that for Delta-specific responses in the Delta breakthrough cohort (δ S1: 0.32, 0.46, 0.63 respectively) (no overlapping 95% confidence intervals) (Figure 4d; Supp Figure 7d). The time of peak for plasma IgG, IgA and Fcγ3aR responses against Omicron BA.2 S1 and ST by the Omicron BA.2 breakthrough cohort (ο BA.2 S1: 14.01, 16.78, 16.95 days respectively) was also much later, taking almost twice as long than that observed with Delta-specific responses in the Delta breakthrough cohort (δ S1: 8.17, 7.69, 7.85 days respectively) (no overlapping 95% confidence intervals) (Figure 4d; Supp Figure 7d).

Finally, comparison of the growth rate and time of peak for IgG, IgA and Fcγ3aR responses between plasma and saliva Omicron BA.2 breakthrough cohort samples identified a trend of notably slower and later Omicron BA.2 spike responses in plasma than in saliva (Figure 4b; Supp Figure 7b). Altogether, the observed poorer growth rates and delays in plasma antibody responses likely account for the overall poorer magnitude of Omicron BA.2 plasma responses detected above two weeks after the Omicron BA.2 breakthrough infections. These differences in antibody kinetics between both humoral compartments highlight the importance of local responses in resolving mucosal infections, particularly in the resolution of Omicron BA.2 breakthrough infections.

## Discussion

Mucosal immunity in the upper respiratory tract is a first line of defense against respiratory infections. Higher levels of salivary antibodies, especially anti-RBD secretory IgA, are associated with protection against breakthrough COVID-19 [3]. Indeed, neutralizing IgA is detected in convalescent COVID-19 individuals, and can recognize a range of RBD mutations [30, 31]. We, and others have previously shown that current IM COVID-19 vaccinations are inefficient in inducing mucosal IgA responses in vaccinated only individuals [1, 5, 6]. However, recent research has highlighted that COVID-19 recovered individuals (convalescent, vaccinated) could induce better salivary IgA responses [2, 4].

Here, we demonstrate that COVID-19 recovered individuals generate robust salivary humoral responses after receiving their first mRNA vaccine, including an enhanced level of salivary IgA. These responses are, however, more biased towards the more conserved regions of the viral spike, namely S2, instead of the more diverse RBD. As such, not surprisingly, COVID-19 recovered individuals only induce slightly better salivary neutralizing antibodies relative to vaccinated only individuals after vaccination. However, importantly, we observed that salivary IgA from COVID-19 recovered individuals had strong and broad cross reactivity across the range of ST from the respective SARS-CoV-2 VoCs. This highlights the importance of cross-reactive mucosal IgA in the first-line of defense against new and emerging SARS-CoV-2 VoCs.

Conversely, unlike after their first IM mRNA vaccination, mucosal IgA responses from COVID-19 recovered individuals were not boosted, but instead decreased after receiving the second IM mRNA vaccine. This suggests that repeated stimulation at the mucosa is required for retaining good site-specific antibody responses and highlights the potential importance of intranasal COVID-19 vaccines in establishing and maintaining mucosal immunity [27, 32]. This also aligns with literature that IM boosters (third dose) were not associated with much additional protection among COVID-19 recovered individuals against Omicron BA.1 infections [33].

Increasingly, studies are suggesting the role of Fc-dependent antibody effector functions in determining the outcome of SARS-CoV-2 infection, particularly in the absence of neutralizing antibodies against emerging COVID-19 variants [34]. Here, we show that COVID-19 recovered individuals also induced better salivary antibody responses capable of engaging FcγRs after two and three antigen exposures, as compared to individuals with only vaccine-induced immunity. This could be due to the retention of tissue-resident memory B cells at the mucosa after COVID-19 [35].

Convalescent individuals who were subsequently vaccinated have been shown to develop broader cross-reactive antibody affinity maturation that can better engage Omicron subvariants than vaccinated only vaccinees [36]. Here, we observed that while a 2^nd^ dose of mRNA COVID-19 ancestral vaccine did induce stronger overall antibody-mediated FcγR engagement in plasma and saliva, it favored responses against the imprinted ancestral wildtype spike. Receiving the bivalent or variant-based vaccines instead of the ancestral vaccine could be more effective in promoting broader cross-reactive antibodies and overcome the limitations of immune imprinting by ancestral SARS-CoV-2 spike [37, 38].

Irrgang et al. recently published that repeated mRNA vaccines drove the responses of non-inflammatory IgG4 in circulation and this class-switching was associated with the reduced capacity for spike-specific antibody-mediated cellular and complement responses [39]. Here, we noted that salivary and tear fluid IgG responses mimicked that found in circulation, particularly in the enrichment of IgG4 responses following the third mRNA vaccination. Additional mRNA boosters may exacerbate subclass restrictions and impact the ability of salivary antibodies to better engage functional responses at the mucosa. As mRNA boosters remain a useful tool in periodically bolstering the humoral responses of vulnerable populations, future studies should evaluate if reduced mRNA vaccine dosages would be more suitable for repeated seasonal boosting of COVID-19 vaccinated populations instead.

COVID-19 vaccination and boosters have been instrumental in reducing disease susceptibility and severity in vaccinated only populations following breakthrough infections, particularly with Omicron VOCs [33]. However, there have been suggestions that ancestral imprinting and antibody feedback from high levels of pre-existing immunity may also restrict humoral responses during breakthrough infections [9-11, 40, 41]. Havervall et al. highlighted that following Omicron BA.1 breakthrough infections, while the rise in nasal IgA responses coincided with the decline in viral load (measured by qPCR), most of the elicited IgA were still targeting the ancestral spike instead of Omicron BA.1 [2].

Here, we demonstrated that pre-existing immunity influences the magnitude of systemic neutralizing responses made following breakthrough infections. Furthermore, a single breakthrough infection appeared insufficient to boost good neutralizing responses at the mucosa in majority of individuals studied, regardless of VoC wave (Delta, Omicron BA.1, Omicron BA.2). This gap in mucosal immunity could still leave individuals recovered from COVID-19 breakthrough infection susceptible to a repeat SARS-CoV-2 infection.

Our findings contrast with observations from “prime and spike” mice models, where a single intranasal booster was sufficient for the robust induction of mucosal antibodies [27]. Nevertheless, despite differences arising from infection and vaccination, our observations do align with human data from the recent Phase I clinical trial where SARS-CoV-2 specific mucosal antibodies were only detectable in a minority of participants receiving intranasal vaccination with ChAdOx1 nCoV-19 [32]. It also remains unclear if the dampened SARS-CoV-2 breakthrough infection resulting from enhanced viral clearance by pre-existing immunity may have limited viral antigen exposure required for generating better variant-specific antibodies. Future studies could address if a repeat SARS-CoV-2 infection or receiving variant-specific vaccines, especially nasal vaccines, after recovering from COVID-19 would enhance mucosal humoral responses.

We also noticed a delay in the time of activation, growth rate and time of peak of variant-specific plasma antibody responses in line with increasing pre-existing immunity within the Omicron BA.2 breakthrough cohorts. It should also be noted that salivary Omicron BA.2-specific responses were quicker to peak than that in plasma. This suggests that local mucosal antibody responses may play a bigger role in the timely control and clearance of mucosal infections arising from emerging COVID-19 variants.

Hybrid immunity has also been commonly used to refer to cohorts of either COVID-19 recovered vaccinees (convalescent, then vaccinated) or individuals with breakthrough infections (vaccinated, then infected). Here, we have demonstrated that while both cohorts are conceptually similar, the order and sites of exposure, as well as their levels of pre-existing immunity can promote different humoral responses, particularly at the mucosa.

Future work could be done to assess how waning levels of pre-existing immunity would impact mucosal responses generated against newer Omicron variants. The impact of repeated mucosal exposures through either acquired infection or receiving intranasal vaccination, particularly with non-ancestral vaccines, on the induction of site-specific antibodies should also be explored. Furthermore, while saliva is a convenient sample for studying mucosal responses, future studies should investigate if mucosal antibody responses, particularly secretory IgA, may be further enriched in nasal fluid instead [42]. We also acknowledge that collecting larger volumes of mucosal samples (saliva, tear fluid, nasal fluid) could allow the future use of cell-based live virus microneutralization assays instead.

## Conclusions

IM COVID-19 vaccinations may be effective in generating systemic immunity to protect against severe disease, but they remain inefficient in eliciting sustained mucosal antibodies, even among COVID-19 recovered individuals. These gaps in mucosal immunity, particularly a lack in mucosal neutralizing antibodies and IgA responses, likely contribute to high rates of breakthrough infections with Omicron variants, highlighting the urgency for effective mucosal COVID-19 vaccines. While pre-existing systemic immunity afforded by current COVID-19 vaccines and boosters facilitate viral clearance, more emphasis should be placed on inducing better local SARS-CoV-2 specific mucosal antibodies.

## Data Availability

All data produced in the present study are available upon reasonable request to the authors

## Acknowledgements

We thank the participants for their generous time and gracious provision of samples. We thank T. Amarasena, R. Esterbauer, K. Wragg, P. Konstandopoulos, G. Gare, K. Field, H. Kelly and A. Kelly (University of Melbourne) for their outstanding technical assistance. We thank G. Taiaroa, P. Kinsella and the molecular staff at the Victorian Infectious Diseases Reference Laboratory for performing rRT-PCR. We thank Dale Godfrey, Nicholas Gherardin and Samuel Redmond for generously sharing their recombinant biotinylated-ACE-2 protein. This study is supported by the Medical Research Future Fund (MRFF) GNT #2005544 to JAJ, SJK, AWC and GNT #2016062 to JAJ, MPD, SJK, AWC. DC, JAJ, MPD, SJK and AWC are supported by NHMRC Investigator grants. The Duke-NUS team is supported by grants from NMRC Singapore (STPRG-FY19-001, COVID19RF-003, and OFLCG19May-0034)

## Supplementary Figure Legends

**Supp Figure 1: Cohort Information and spike schematic**

Details of the pre-pandemic controls, vaccinated only vaccinees and COVID-19 recovered vaccinees included in the study **(a)**. Details of vaccination status and samples collected for the COVID-19 breakthrough infection cohorts post symptom onset **(b, c)**. Circles depict the timepoints where paired plasma and saliva samples were collected in the presence (open circles) or absence (closed circles) of a nasal swab sample. Schematic of the SARS-CoV-2 spike protein describing the four different types of spike proteins used in the multiplex assays **(d)**.

**Supp Figure 2: Titration curves for SARS-CoV-2 spike coupled arrays**

Titration curves depict the plasma and salivary total IgG and IgA responses against ancestral wildtype spike antigens using samples from three vaccinated individuals at the respective timepoints **(a)**. Titration curves depict the plasma and salivary total IgG and IgA responses against ancestral wildtype spike and the VoC spikes using pooled plasma from COVID-19 boostered individuals and the saliva sample from a boostered individual who seroconverted strongly during their delta breakthrough infection **(b)**. Asterisks define the respective dilutions chosen for the multiplex assays.

**Supp Figure 3: Comparisons of mucosal antibody responses between vaccinated only and COVID-19 recovered vaccinees after 2 and 3 antigen exposures**

Bar graphs describe the key differences in ancestral SARS-CoV-2 spike-specific salivary **(a, b)** antibody responses between vaccinated only and COVID-19 recovered cohorts after two **(a)** and three **(b)** antigens exposures. Bar graphs describing key features in ancestral SARS-CoV-2 spike-specific tear antibody responses between vaccinated only and COVID-19 recovered cohorts after three antigens exposures **(c)**. Statistical significance was calculated using the two-tailed Mann-Whitney *U* test. Bar graphs also illustrate the changes in anti-Spike 2 IgA responses in saliva **(d)** and tear fluid **(e)** of COVID-19 recovered individuals after their first and second mRNA vaccination. Statistical significance was calculated using Friedman’s test followed by Dunn’s test for multiple comparisons. Where significant or trending significance, *p*-values were reported (* p≤0.05; ** p≤0.01; *** p≤0.001; **** p≤0.0001). Spearman correlation of salivary IgG2 and IgG4 responses against FcγR engagement in the vaccinated only cohort after two **(f)** or three **(g)** mRNA vaccinations.

**Supp Figure 4: Plasma antibodies from COVID-19 recovered vaccinees show strong IgG2 and IgG4 responses after multiple COVID-19 mRNA vaccination.**

Plasma antibody isotype and subclass responses from both cohorts against the various SARS-CoV-2 spike antigens were compiled into respective radar plots **(a)**. The individual median antibody isotype/subclass response for each spike antigen was transformed into percentages using the antigen-specific MFI from the 98^th^ percentile for that detector (98^th^ percentile was chosen to minimize the impact of outliers on the data transformation). PCA of all 40 antibody features for vaccinated only (closed circles) and COVID-19 recovered (open squares) individuals after two **(b)** and three **(e)** antigen exposures. Loading plots and bar graphs describe the key differences between both cohorts after two **(c, d)** and three **(f, g)** antigens exposure. Statistical significance was calculated using the two-tailed Mann-Whitney *U* test and where significant or trending significance, *p*-values were reported (* p≤0.05; ** p≤0.01; *** p≤0.001; **** p≤0.0001). Spearman correlation of plasma IgG2 and IgG4 responses against FcγR engagement in the vaccinated only cohort after two **(e)** or three **(f)** mRNA vaccinations..

**Supp Figure 5: No changes in salivary or plasma IgG4 responses after repeated adenoviral vector COVID-19 vaccination.**

Paired saliva and plasma samples were collected pre- and post-vaccination from vaccinated only vaccinees at the indicated time-points **(a)**. Bar graphs illustrate the changes in total IgG and IgG4 responses against Spike 1 and Spike 2 respectively in saliva **(b)** and plasma **(c)** of vaccinated only vaccinees after their second ChAdOx nCoV-19 vaccine (orange) or mRNA booster (green). Statistical significance was calculated using Friedman’s test followed by Dunn’s test for multiple comparisons. Where significant or trending significance, *p*-values were reported (* p≤0.05; ** p≤0.01; *** p≤0.001; **** p≤0.0001).

**Supp Figure 6: Viral load kinetics of breakthrough infections and influence of pre-existing vaccine-induced immunity on elicited antibody responses**

Modelled kinetic curves describe the viral load (Ct value) post-symptom onset as determined by nasal swab samples collected during the Delta **(a)** (Red) and Omicron BA.2 **(c)** (Green) COVID-19 breakthrough waves. Heat maps compare the salivary **(c)** and plasma **(e)** inhibition of RBD-ACE2 interactions against the ancestral wildtype (WT) SARS-CoV-2 or the VoCs (α, Alpha; δ, Delta; β, Beta; ο BA.1, Omicron BA.1; ο BA.2, Omicron BA.2) between the start and clearance of COVID-19 breakthrough infections. Heat maps also describe the salivary **(d)** and plasma **(f)** antibody responses capable of FcγR3a engagement that are elicited early and 2 weeks post-symptom onset of COVID-19 breakthrough infections. Significant differences between both timepoints were calculated using Friedman’s test followed by Dunn’s test for multiple comparisons. Spearman correlation of plasma **(g, h)** and saliva **(i, j)** IgG (open circles) and IgA (closed squares) responses against viral load (Ct value) in the Delta **(g, i)** (Red) and Omicron BA.2 **(h, j)** (Green) COVID-19 breakthrough waves. Where significant, *p*-values were reported (* p≤0.05; ** p≤0.01; *** p≤0.001; **** p≤0.0001).

**Supp Figure 7: Pre-existing vaccine-induced immunity modulate antibody kinetics against spike trimer during COVID-19 breakthrough infection**

Modelled kinetic curves (WT: black; Delta: red; Omicron BA.1: blue; Omicron BA.2: green) describe the ancestral wildtype and variant-specific Spike Trimer antibody responses from serially collected saliva **(a)** and plasma **(c)** samples during Delta or Omicron BA.2 breakthrough infection for up to 40 days post-symptom onset. Connected dotted lines indicate serial samples from the same individual. Lines with open circles at the bottom of each graph reflect samples that were excluded from the model for being below the threshold of detection (2 S.D. background readings). Dot plots displaying 95% confidence intervals beside each row of kinetic curves list the calculated growth rate, time to activation and time to peak of variant-specific salivary **(b)** and plasma **(d)** responses (Delta: red; Omicron BA.2: green) by their respective breakthrough cohorts (eg: Delta variant responses during the Delta breakthroughs).

## Supplementary Tables

**Supp Table 8: Cohort information for pre-pandemic controls and COVID-19 vaccinees**

**Supp Table 9: Cohort information for individuals with COVID-19 breakthrough infections**

**Supp Table 10: Parameters and confidence intervals for modelled kinetic curves for COVID-19 breakthrough infections**

