## Supplementary Figures for "Lasting first impression: Pre-existing immunity restricts mucosal antibody responses during Omicron breakthrough"

**a)** Summary of pre-pandemic controls and vaccinated cohorts

| | Pre-pandemic Controls | Comirnaty / BNT162b2 (Pfizer-BioNtech) vaccinees | | ChAdOx1 nCoV-19 (AstraZeneca) vaccinees |
|---|---|---|---|---|
| **Variables** | Pre-pandemic (n=20) | Vaccinated only (n=20) | COVID-19 Recovered (Convalescent, vaccinated) (n=10) | Vaccinated only (n=17) |
| **Age, mean (range), years** | 30.5 (21 – 60) | 34.6 (26 – 58) | 49.5 (24 – 65) | 46.5 (23 – 69) |
| **Gender** | | | | |
| **Female (%)** | 16 (80.0%) | 13 (65.0%) | 6 (60.0%) | 10 (58.8) |
| **Male (%)** | 4 (20.0%) | 7 (35.0%) | 4 (40.0%) | 7 (41.2) |
| **Time from symptom onset till sample collection, mean (range), days** | | | 478.1 (412 – 534) | |
| **Vaccination** | | | | |
| **Time after 1st shot till sample collection, mean (range), days** | | 9.1 (6 – 12) | 12.5 (7 – 18) | 13.4 (7 – 22) |
| **Time between 1st and 2nd shot, mean (range), days** | | 24.0 (21 – 33) | 28.0 (21 – 48) | 83 (70 – 92) |
| **Time after 2nd shot till sample collection, mean (range), days** | | 14.1 (12 – 22) | 20.1 (10 – 34) | 13.4 (7 – 22) |
| **Time between 2nd shot and 5 months sample collection, mean (range), days** | | 218.8 (188 – 226) | | 149.6 (101 – 184) |
| **Time after 3rd shot till sample collection, mean (range), days** | | 12.4 (10 – 15) | | 20.1 (10 – 31) |

**b)** Summary of breakthrough cohorts

| | Delta breakthroughs | Omicron BA.2 breakthroughs |
|---|---|---|
| **Variables** | (n=8) | (n=10) |
| **Age, mean (range), years** | 34.6 (19 – 58) | 54.3 (40 – 66) |
| **Gender** | | |
| **Female (%)** | 5 (62.5%) | 5 (50.0%) |
| **Male (%)** | 3 (37.5%) | 5 (50.0%) |
| **Vaccination doses prior to breakthrough infection (%)** | 2 (100.0%) | 3 (100.0%) |
| **Time from last vaccination till breakthrough infection, mean (range), months** | 3.3 (0.3 – 5.3) | 3.2 (1.6 – 5.3) |

**c)** Sampling timepoints for breakthrough infections

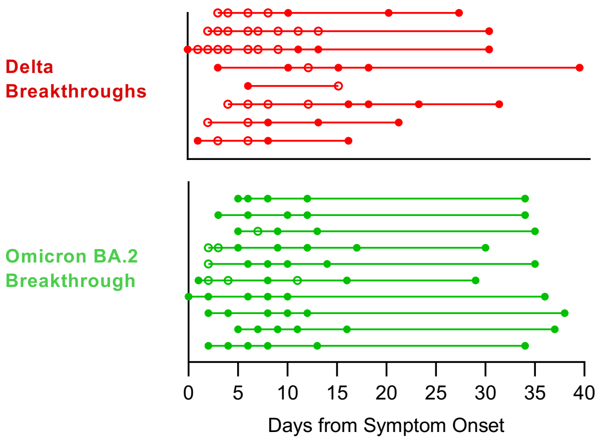


**d)** Schematic of SARS-CoV-2 spike protein

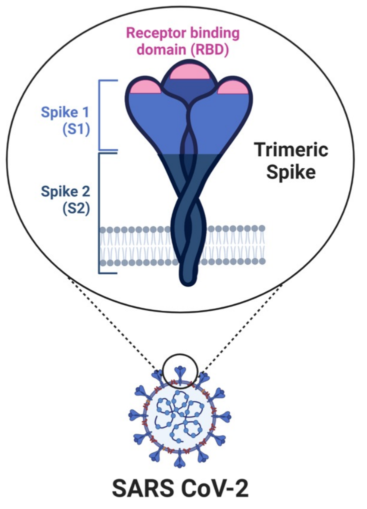

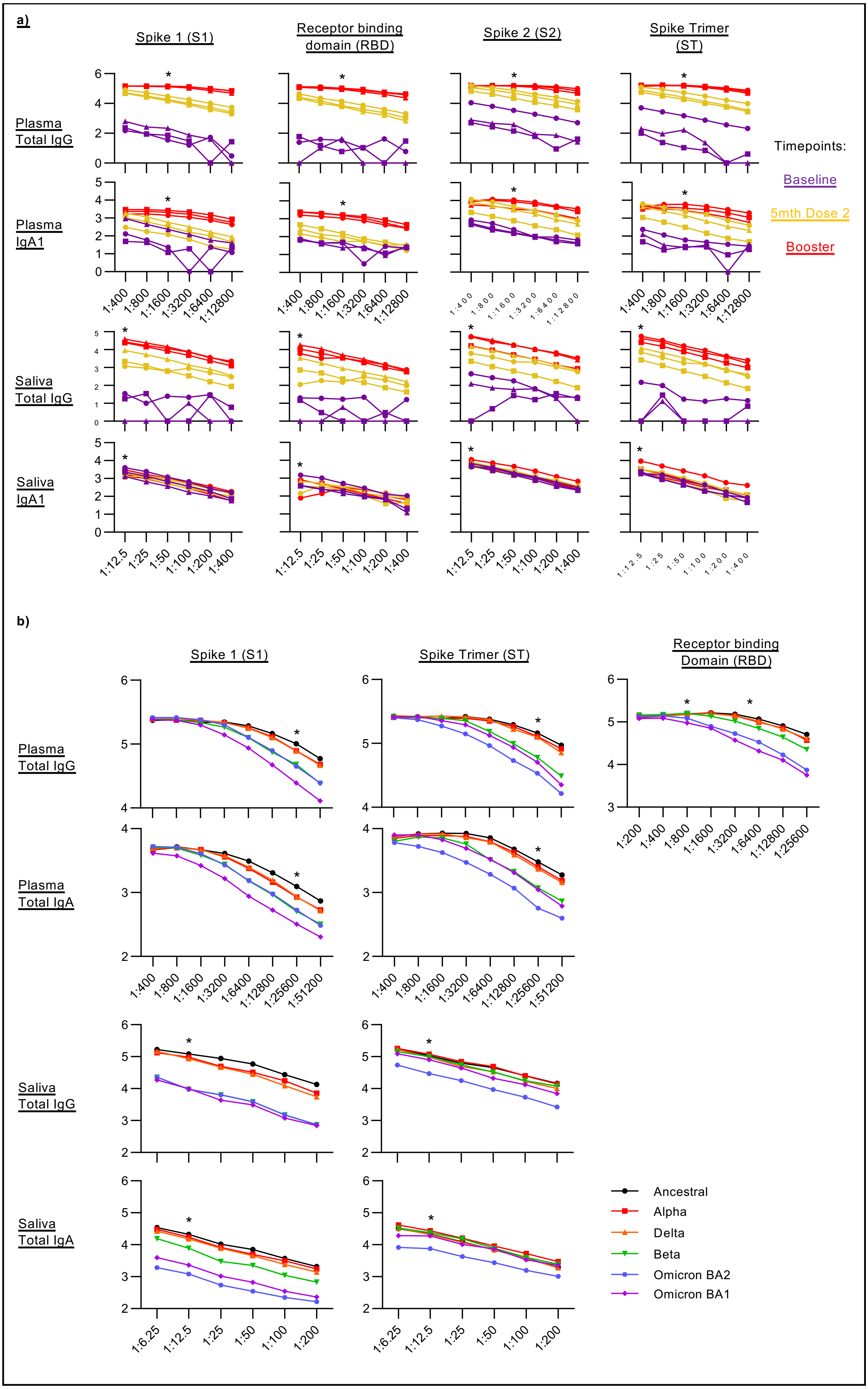

a)
Spike 1 (S1)
Receptor binding domain (RBD)
Spike 2 (S2)
Spike Trimer (ST)
Plasma Total IgG
Plasma IgA1
Saliva Total IgG
Saliva IgA1
Timepoints:
Baseline
5mth Dose 2
Booster
1:400 1:800 1:1600 1:3200 1:6400 1:12800
1:12.5 1:25 1:50 1:100 1:200 1:400
b)
Spike 1 (S1)
Spike Trimer (ST)
Receptor binding Domain (RBD)
Plasma Total IgG
Plasma Total IgA
Saliva Total IgG
Saliva Total IgA
1:400 1:800 1:1600 1:3200 1:6400 1:12800 1:25600 1:51200
1:200 1:400 1:800 1:1600 1:3200 1:6400 1:12800 1:25600
1:6.25 1:12.5 1:25 1:50 1:100 1:200
Ancestral
Alpha
Delta
Beta
Omicron BA2
Omicron BA1

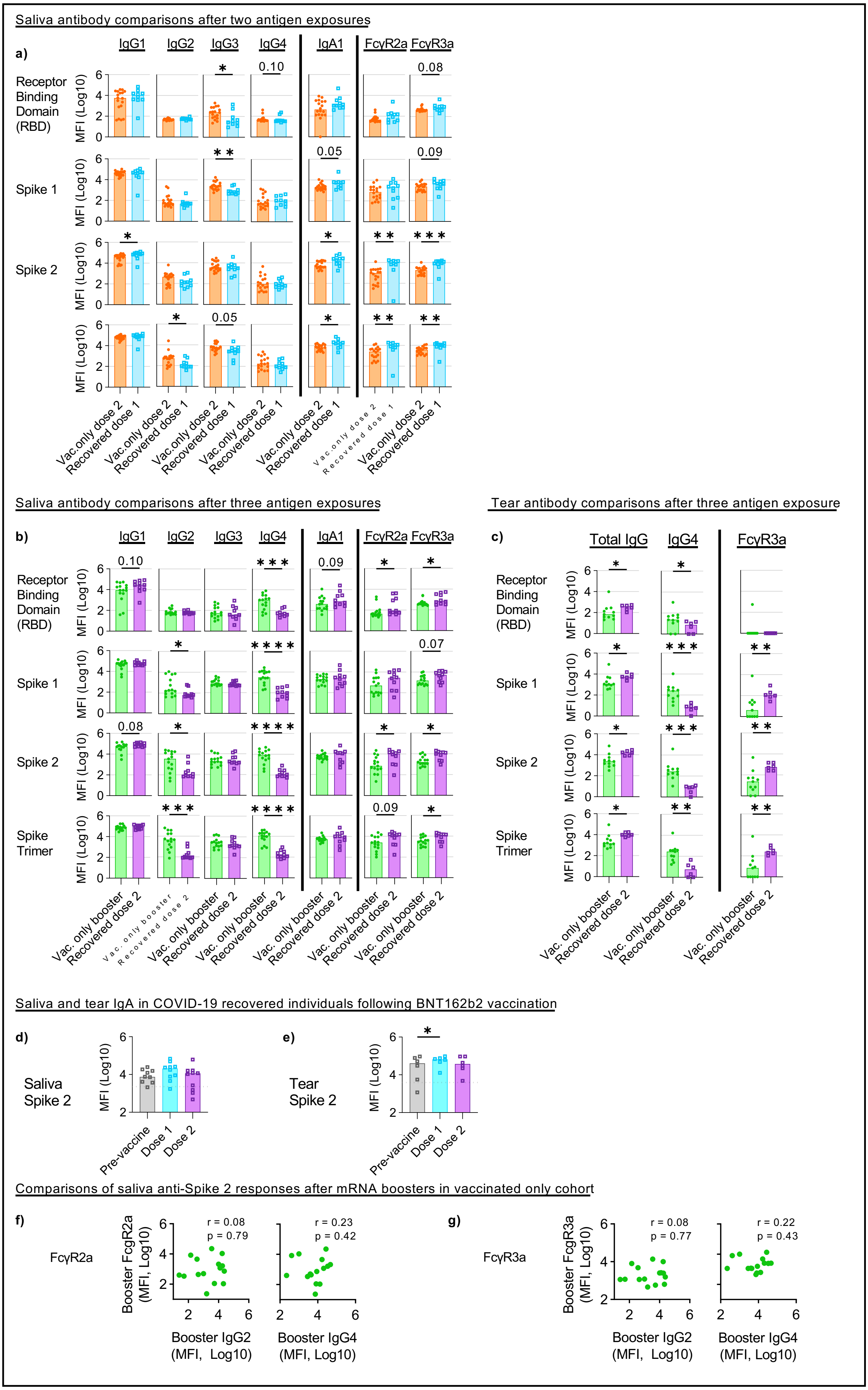
Saliva antibody comparisons after two antigen exposures
a)
IgG1 IgG2 IgG3 IgG4 IgA1 FcγR2a FcγR3a
Receptor Binding Domain (RBD)
Spike 1
Spike 2
MFI (Log10)
Vac.only dose 2
Recovered dose 1
Saliva antibody comparisons after three antigen exposures
b)
Spike Trimer
Vac. only booster
Recovered dose 2
Tear antibody comparisons after three antigen exposure
c)
Total IgG IgG4 FcγR3a
Saliva and tear IgA in COVID-19 recovered individuals following BNT162b2 vaccination
d)
Saliva Spike 2
e)
Tear Spike 2
Pre-vaccine
Dose 1
Dose 2
Comparisons of saliva anti-Spike 2 responses after mRNA boosters in vaccinated only cohort
f)
FcγR2a
Booster FcgR2a (MFI, Log10)
r = 0.08 p = 0.79
r = 0.23 p = 0.42
Booster IgG2 (MFI, Log10)
Booster IgG4 (MFI, Log10)
g)
FcγR3a
Booster FcgR3a (MFI, Log10)
r = 0.08 p = 0.77
r = 0.22 p = 0.43

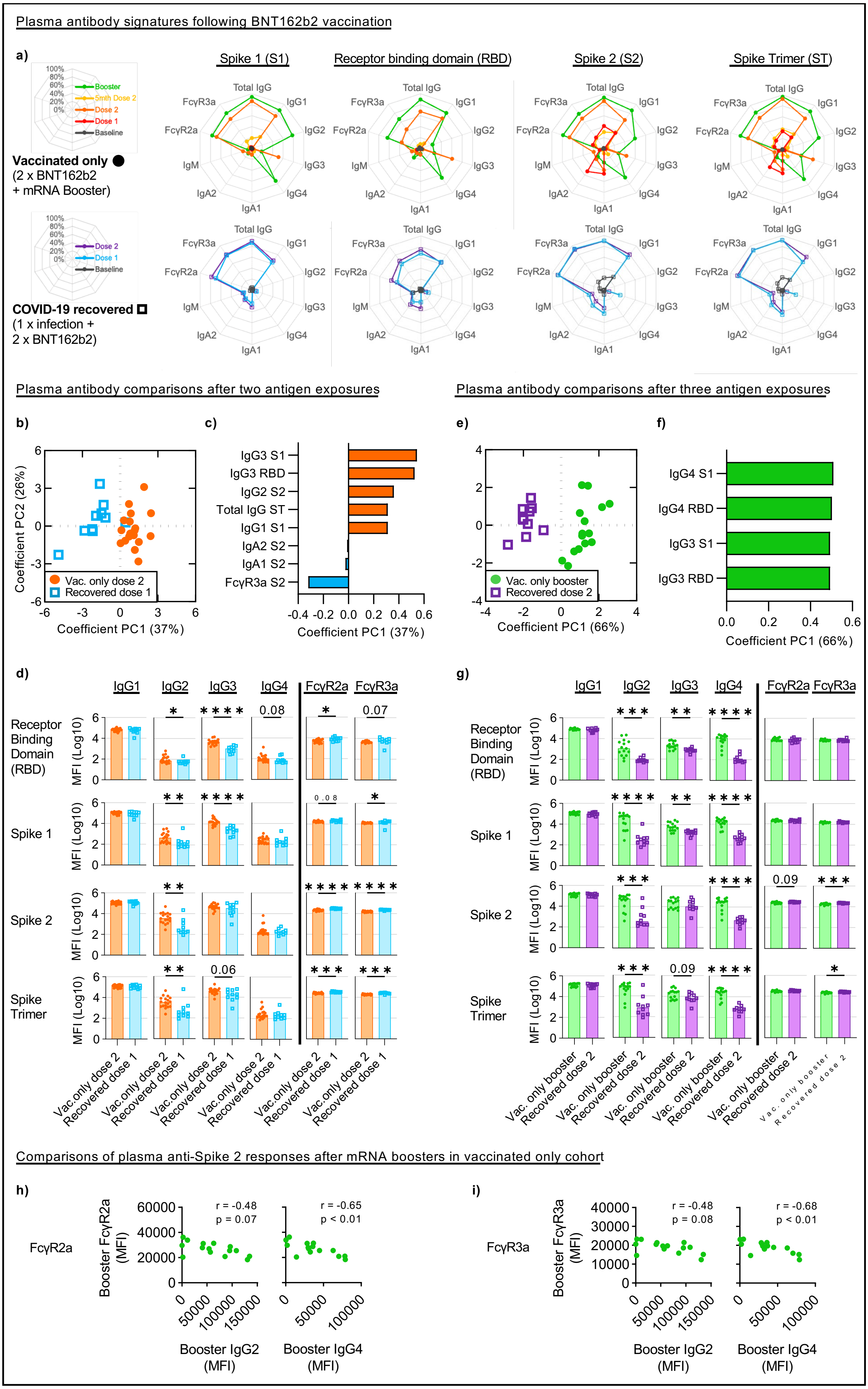

Plasma antibody signatures following BNT162b2 vaccination
a)
Booster
5mth Dose 2
Dose 2
Dose 1
Baseline
Vaccinated only
(2 x BNT162b2
+ mRNA Booster)
Dose 2
Dose 1
Baseline
COVID-19 recovered
(1 x infection +
2 x BNT162b2)
Spike 1 (S1)
Receptor binding domain (RBD)
Spike 2 (S2)
Spike Trimer (ST)
Total IgG
IgG1
IgG2
IgG3
IgG4
IgA1
IgA2
IgM
FcγR2a
FcγR3a
Plasma antibody comparisons after two antigen exposures
Plasma antibody comparisons after three antigen exposures
b)
Coefficient PC2 (26%)
Coefficient PC1 (37%)
Vac. only dose 2
Recovered dose 1
c)
IgG3 S1
IgG3 RBD
IgG2 S2
Total IgG ST
IgG1 S1
IgA2 S2
IgA1 S2
FcγR3a S2
Coefficient PC1 (37%)
e)
Coefficient PC1 (66%)
Vac. only booster
Recovered dose 2
f)
IgG4 S1
IgG4 RBD
IgG3 S1
IgG3 RBD
Coefficient PC1 (66%)
d)
Receptor Binding Domain (RBD)
Spike 1
Spike 2
Spike Trimer
MFI (Log10)
Vac.only dose 2
Recovered dose 1
g)
Vac. only booster
Recovered dose 2
Comparisons of plasma anti-Spike 2 responses after mRNA boosters in vaccinated only cohort
h)
FcγR2a
Booster FcγR2a (MFI)
r = -0.48
p = 0.07
r = -0.65
p < 0.01
Booster IgG2 (MFI)
Booster IgG4 (MFI)
i)
FcγR3a
Booster FcγR3a (MFI)
r = -0.48
p = 0.08
r = -0.68
p < 0.01
Booster IgG2 (MFI)
Booster IgG4 (MFI)

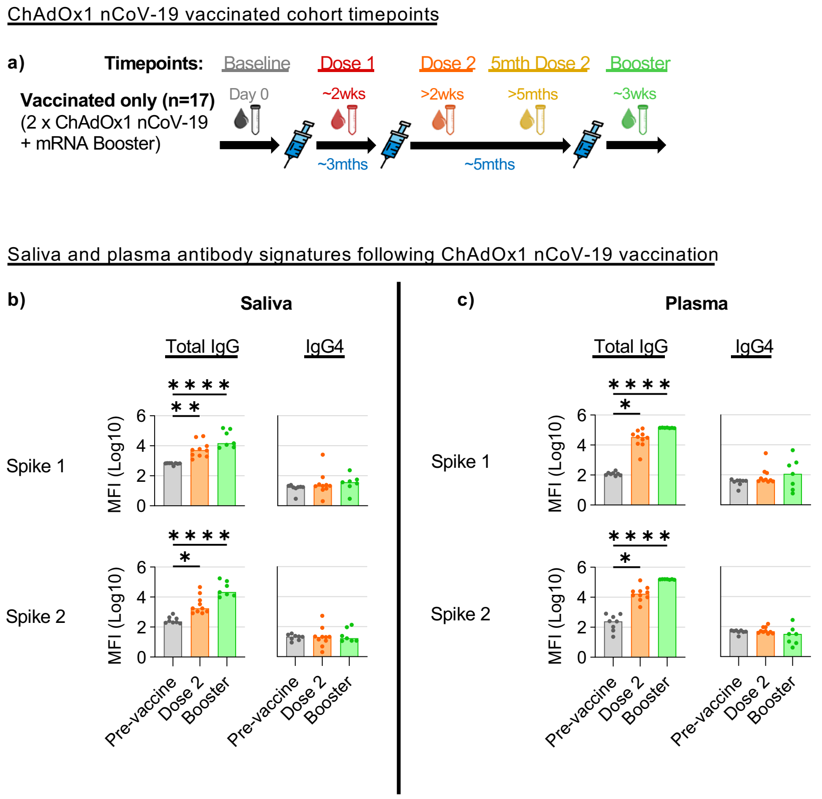
ChAdOx1 nCoV-19 vaccinated cohort timepoints
a)
Timepoints:
Baseline
Dose 1
Dose 2
5mth Dose 2
Booster
Vaccinated only (n=17)
(2 x ChAdOx1 nCoV-19
+ mRNA Booster)
Day 0
~2wks
>2wks
>5mths
~3wks
~3mths
~5mths
Saliva and plasma antibody signatures following ChAdOx1 nCoV-19 vaccination
b)
Saliva
c)
Plasma
Total IgG
IgG4
Spike 1
Spike 2
MFI (Log10)
Pre-vaccine
Dose 2
Booster

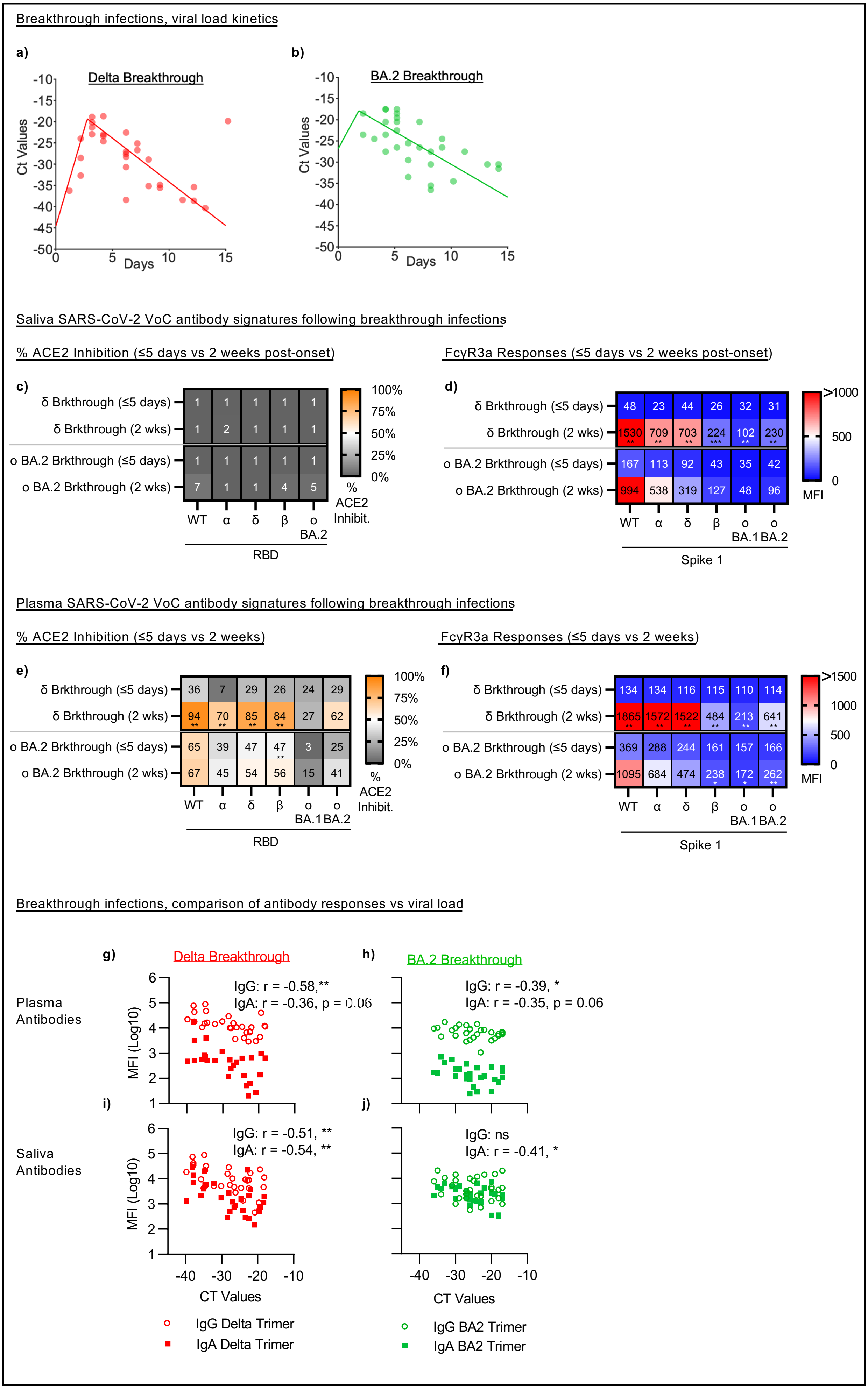
Breakthrough infections, viral load kinetics
a)
Delta Breakthrough
Ct Values
Days
b)
BA.2 Breakthrough
Ct Values
Days
Saliva SARS-CoV-2 VoC antibody signatures following breakthrough infections
% ACE2 Inhibition (≤5 days vs 2 weeks post-onset)
c)
δ Brkthrough (≤5 days) 1 1 1 1 1
δ Brkthrough (2 wks) 1 2 1 1 1
ο BA.2 Brkthrough (≤5 days) 1 1 1 1 1
ο BA.2 Brkthrough (2 wks) 7 1 1 4 5
WT α δ β ο BA.2
RBD
100% 75% 50% 25% 0%
% ACE2 Inhibit.
FcγR3a Responses (≤5 days vs 2 weeks post-onset)
d)
δ Brkthrough (≤5 days) 48 23 44 26 32 31
δ Brkthrough (2 wks) 1530 709 703 224 102 230
ο BA.2 Brkthrough (≤5 days) 167 113 92 43 35 42
ο BA.2 Brkthrough (2 wks) 994 538 319 127 48 96
WT α δ β ο BA.1 ο BA.2
Spike 1
>1000 500 0
MFI
Plasma SARS-CoV-2 VoC antibody signatures following breakthrough infections
% ACE2 Inhibition (≤5 days vs 2 weeks)
e)
δ Brkthrough (≤5 days) 36 7 29 26 24 29
δ Brkthrough (2 wks) 94 70 85 84 27 62
ο BA.2 Brkthrough (≤5 days) 65 39 47 47 3 25
ο BA.2 Brkthrough (2 wks) 67 45 54 56 15 41
WT α δ β ο BA.1 ο BA.2
RBD
100% 75% 50% 25% 0%
% ACE2 Inhibit.
FcγR3a Responses (≤5 days vs 2 weeks)
f)
δ Brkthrough (≤5 days) 134 134 116 115 110 114
δ Brkthrough (2 wks) 1865 1572 1522 484 213 641
ο BA.2 Brkthrough (≤5 days) 369 288 244 161 157 166
ο BA.2 Brkthrough (2 wks) 1095 684 474 238 172 262
WT α δ β ο BA.1 ο BA.2
Spike 1
>1500 1000 500 0
MFI
Breakthrough infections, comparison of antibody responses vs viral load
g)
Delta Breakthrough
Plasma Antibodies
IgG: r = -0.58,**
IgA: r = -0.36, p = 0.06
MFI (Log10)
h)
BA.2 Breakthrough
IgG: r = -0.39, *
IgA: r = -0.35, p = 0.06
i)
Saliva Antibodies
IgG: r = -0.51, **
IgA: r = -0.54, **
MFI (Log10)
j)
IgG: ns
IgA: r = -0.41, *
CT Values
CT Values
IgG Delta Trimer
IgA Delta Trimer
IgG BA2 Trimer
IgA BA2 Trimer

Saliva antibody responses over time, breakthrough infections

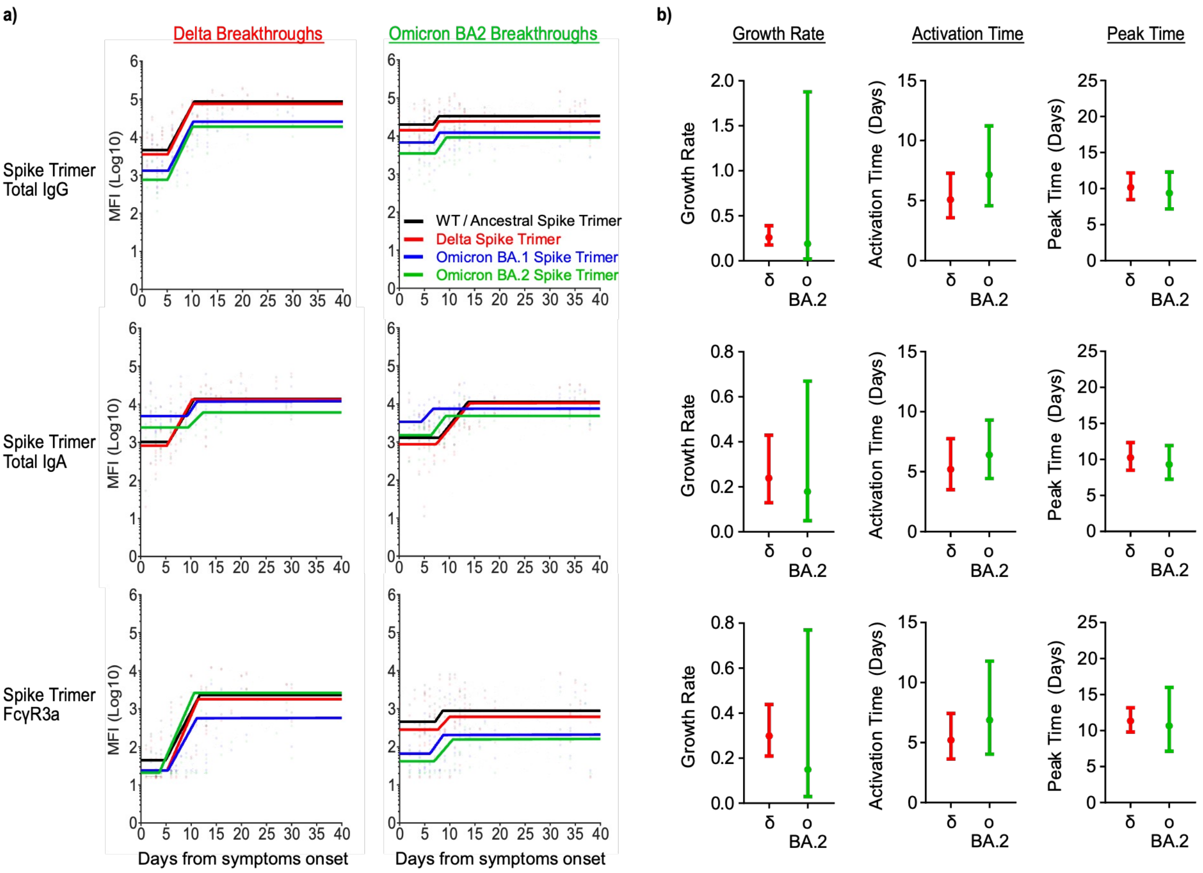


Plasma antibody responses over time, breakthrough infections

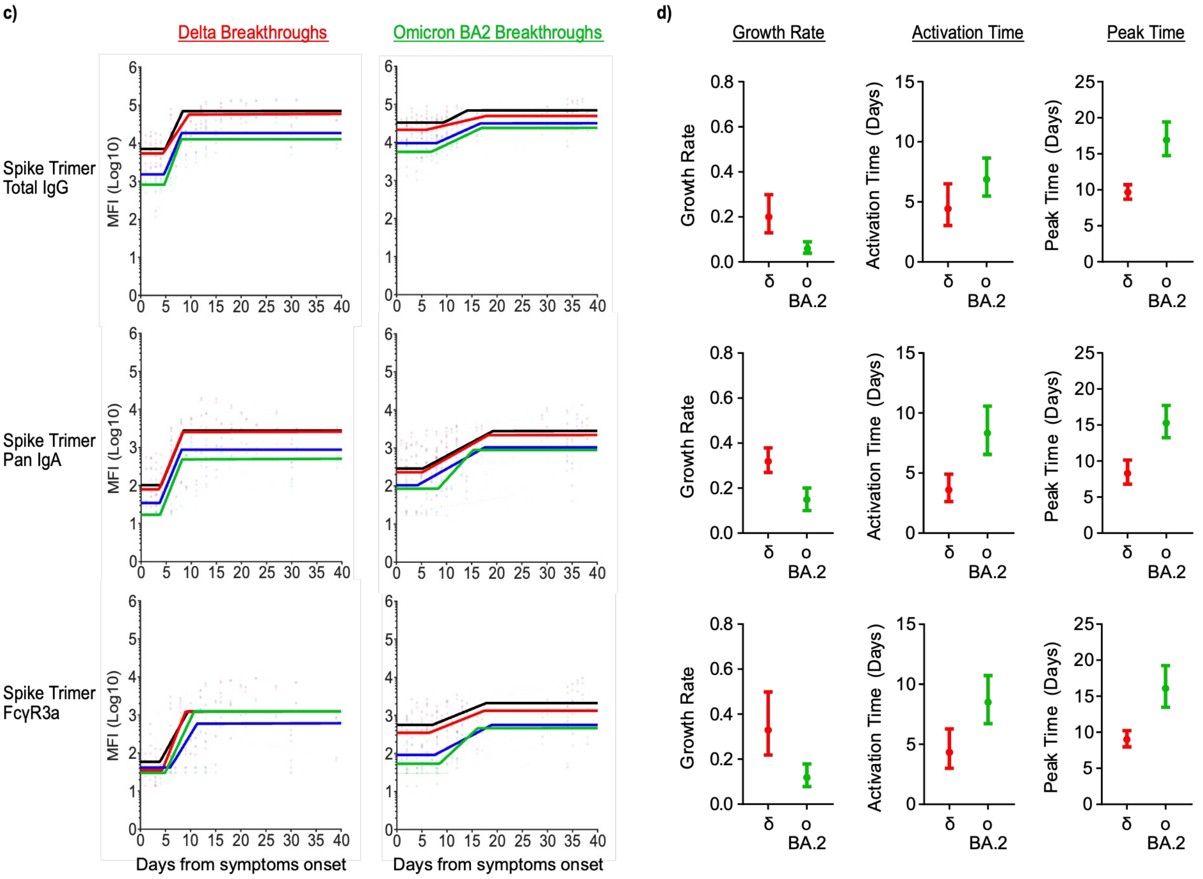
