## Supplementary Table 8 for "Lasting first impression: Pre-existing immunity restricts mucosal antibody responses during Omicron breakthrough"

Supplementary Table 8: Cohort information for COVID-19 vaccinees (V) and pre-pandemic controls (P)

| Donor: | Gender: | Age: | Time from  previous  COVID-19  (days): | 1st  vaccine  dose: | Timepoint  post 1st dose  (days): | Time  between 1st and 2nd dose: | 2nd  vaccine dose: | Timepoint  post 2nd dose  (days): | Extended  timepoint  post  2nd dose  (days) | 3rd vaccine dose: | Timepoint  post 3rd dose  (days): |
| --- | --- | --- | --- | --- | --- | --- | --- | --- | --- | --- | --- |
| V01 | F | In their 50s | n.d. | BNT162b2 | n.d. | 22 | BNT162b2 | 12 | 224 | BNT162b2 | 11 |
| V02 | M | In their 30s | n.d. | BNT162b2 | 11 | 23 | BNT162b2 | 13 | 225 | mRNA-1273 | 11 |
| V03 | F | In their 20s | n.d. | BNT162b2 | 10 | 22 | BNT162b2 | 13 | n.d. | n.d. | n.d. |
| V04 | F | In their 20s | n.d. | BNT162b2 | 9 | 21 | BNT162b2 | 13 | 225 | BNT162b2 | 14 |
| V05 | M | In their 30s | n.d. | BNT162b2 | n.d. | 27 | BNT162b2 | 14 | 219 | BNT162b2 | 11 |
| V06 | F | In their 30s | n.d. | BNT162b2 | 9 | 28 | BNT162b2 | 13 | n.d. | n.d. | n.d. |
| V07 | F | In their 30s | n.d. | BNT162b2 | 8 | 21 | BNT162b2 | 12 | 224 | BNT162b2 | 13 |
| V08 | F | In their 30s | n.d. | BNT162b2 | n.d. | 29 | BNT162b2 | 13 | 217 | BNT162b2 | 15 |
| V09 | F | In their 20s | n.d. | BNT162b2 | 10 | 24 | BNT162b2 | 18 | 223 | BNT162b2 | 10 |
| V10 | F | In their 20s | n.d. | BNT162b2 | n.d. | 33 | BNT162b2 | 14 | 213 | BNT162b2 | 14 |
| V11 | M | In their 30s | n.d. | BNT162b2 | 10 | 22 | BNT162b2 | 13 | 225 | BNT162b2 | 10 |
| V12 | F | In their 30s | n.d. | BNT162b2 | 10 | 22 | BNT162b2 | 13 | 225 | BNT162b2 | 13 |
| V13 | M | In their 30s | n.d. | BNT162b2 | 6 | 23 | BNT162b2 | 15 | n.d. | n.d. | n.d. |
| V14 | F | In their 30s | n.d. | BNT162b2 | 6 | 23 | BNT162b2 | 15 | 220 | BNT162b2 | 14 |
| V15 | M | In their 30s | n.d. | BNT162b2 | n.d. | 21 | BNT162b2 | 13 | 225 | BNT162b2 | 13 |
| V16 | F | In their 50s | n.d. | BNT162b2 | 7 | 21 | BNT162b2 | 13 | 225 | mRNA-1273 | 11 |
| V17 | M | In their 30s | n.d. | BNT162b2 | 10 | 21 | BNT162b2 | 14 | 226 | BNT162b2 | 13 |
| V18 | M | In their 40s | n.d. | BNT162b2 | 12 | 33 | BNT162b2 | 22 | n.d. | n.d. | n.d. |
| V19 | F | In their 30s | n.d. | BNT162b2 | n.d. | 23 | BNT162b2 | n.d. | 197 | BNT162b2 | 15 |
| V20 | F | In their 30s | n.d. | BNT162b2 | n.d. | 21 | BNT162b2 | n.d. | 188 | BNT162b2 | 10 |
| V21 | F | In their 20s | n.d. | ChAdOx nCoV-19 | 15 | 85 | ChAdOx nCoV-19 | 14 | 168 | mRNA-1273 | 14 |
| V22 | F | In their 50s | n.d. | ChAdOx nCoV-19 | 13 | 85 | ChAdOx nCoV-19 | 12 | n.d. | n.d. | n.d. |
| V23 | M | In their 50s | n.d. | ChAdOx nCoV-19 | 13 | 80 | ChAdOx nCoV-19 | 17 | n.d. | n.d. | n.d. |
| V24 | F | In their 60s | n.d. | ChAdOx nCoV-19 | 22 | 84 | ChAdOx nCoV-19 | 14 | n.d. | n.d. | n.d. |
| V25 | M | In their 60s | n.d. | ChAdOx nCoV-19 | 14 | 70 | ChAdOx nCoV-19 | 7 | n.d. | n.d. | n.d. |
| V26 | M | In their 60s | n.d. | ChAdOx nCoV-19 | 11 | 84 | ChAdOx nCoV-19 | 10 | n.d. | n.d. | n.d. |
| V27 | M | In their 20s | n.d. | ChAdOx nCoV-19 | 15 | 71 | ChAdOx nCoV-19 | 22 | n.d. | n.d. | n.d. |
| V28 | M | In their 50s | n.d. | ChAdOx nCoV-19 | 13 | 81 | ChAdOx nCoV-19 | n.d. | n.d. | n.d. | n.d. |
| V29 | M | In their 60s | n.d. | ChAdOx nCoV-19 | 7 | 84 | ChAdOx nCoV-19 | 7 | n.d. | n.d. | n.d. |
| V30 | M | In their 50s | n.d. | ChAdOx nCoV-19 | 16 | 84 | ChAdOx nCoV-19 | 16 | n.d. | n.d. | n.d. |
| V31 | F | In their 50s | n.d. | ChAdOx nCoV-19 | 8 | 84 | ChAdOx nCoV-19 | 15 | n.d. | n.d. | n.d. |
| V32 | F | In their 30s | n.d. | ChAdOx nCoV-19 | n.d. | 81 | ChAdOx nCoV-19 | n.d. | 101 | BNT162b2 | 14 |
| V33 | F | In their 20s | n.d. | ChAdOx nCoV-19 | n.d. | 84 | ChAdOx nCoV-19 | n.d. | 124 | BNT162b2 | 10 |
| V34 | F | In their 20s | n.d. | ChAdOx nCoV-19 | n.d. | 84 | ChAdOx nCoV-19 | n.d. | 134 | BNT162b2 | 14 |
| V35 | F | In their 50s | n.d. | ChAdOx nCoV-19 | n.d. | 91 | ChAdOx nCoV-19 | n.d. | 158 | BNT162b2 | 29 |
| V36 | F | In their 30s | n.d. | ChAdOx nCoV-19 | n.d. | 90 | ChAdOx nCoV-19 | n.d. | 178 | BNT162b2 | 29 |
| V37 | F | In their 20s | n.d. | ChAdOx nCoV-19 | n.d. | 92 | ChAdOx nCoV-19 | n.d. | 184 | BNT162b2 | 31 |
| V38 | F | In their 50s | 501 | BNT162b2 | 18 | 22 | BNT162b2 | 19 | n.d. | n.d. | n.d. |
| V39 | M | In their 60s | 412 | BNT162b2 | 7 | 48 | BNT162b2 | 22 | n.d. | n.d. | n.d. |
| V40 | F | In their 20s | 534 | BNT162b2 | 7 | 25 | BNT162b2 | 19 | n.d. | n.d. | n.d. |
| V41 | M | In their 50s | 483 | BNT162b2 | 12 | 31 | BNT162b2 | 16 | n.d. | n.d. | n.d. |
| V42 | F | In their 60s | 479 | BNT162b2 | 12 | 21 | BNT162b2 | 13 | n.d. | n.d. | n.d. |
| V43 | M | In their 60s | 493 | BNT162b2 | 13 | 21 | BNT162b2 | 34 | n.d. | n.d. | n.d. |
| V44 | F | In their 50s | 493 | BNT162b2 | 13 | 21 | BNT162b2 | 34 | n.d. | n.d. | n.d. |
| V45 | F | In their 20s | 431 | BNT162b2 | 13 | 25 | BNT162b2 | 13 | n.d. | n.d. | n.d. |
| V46 | M | In their 50s | 438 | BNT162b2 | 14 | 21 | BNT162b2 | 10 | n.d. | n.d. | n.d. |
| V47 | F | In their 30s | 517 | BNT162b2 | 16 | 42 | BNT162b2 | 21 | n.d. | n.d. | n.d. |
| P01 | F | In their 30s | n.d. | n.d. | n.d. | n.d. | n.d. | n.d. | n.d. | n.d. | n.d. |
| P02 | F | In their 20s | n.d. | n.d. | n.d. | n.d. | n.d. | n.d. | n.d. | n.d. | n.d. |
| P03 | F | In their 30s | n.d. | n.d. | n.d. | n.d. | n.d. | n.d. | n.d. | n.d. | n.d. |
| P04 | M | In their 20s | n.d. | n.d. | n.d. | n.d. | n.d. | n.d. | n.d. | n.d. | n.d. |
| P05 | F | In their 30s | n.d. | n.d. | n.d. | n.d. | n.d. | n.d. | n.d. | n.d. | n.d. |
| P06 | F | In their 20s | n.d. | n.d. | n.d. | n.d. | n.d. | n.d. | n.d. | n.d. | n.d. |
| P07 | F | In their 20s | n.d. | n.d. | n.d. | n.d. | n.d. | n.d. | n.d. | n.d. | n.d. |
| P08 | F | In their 20s | n.d. | n.d. | n.d. | n.d. | n.d. | n.d. | n.d. | n.d. | n.d. |
| P09 | F | In their 30s | n.d. | n.d. | n.d. | n.d. | n.d. | n.d. | n.d. | n.d. | n.d. |
| P10 | F | In their 30s | n.d. | n.d. | n.d. | n.d. | n.d. | n.d. | n.d. | n.d. | n.d. |
| P11 | M | In their 30s | n.d. | n.d. | n.d. | n.d. | n.d. | n.d. | n.d. | n.d. | n.d. |
| P12 | F | In their 20s | n.d. | n.d. | n.d. | n.d. | n.d. | n.d. | n.d. | n.d. | n.d. |
| P13 | M | In their 30s | n.d. | n.d. | n.d. | n.d. | n.d. | n.d. | n.d. | n.d. | n.d. |
| P14 | F | In their 30s | n.d. | n.d. | n.d. | n.d. | n.d. | n.d. | n.d. | n.d. | n.d. |
| P15 | F | In their 30s | n.d. | n.d. | n.d. | n.d. | n.d. | n.d. | n.d. | n.d. | n.d. |
| P16 | F | In their 20s | n.d. | n.d. | n.d. | n.d. | n.d. | n.d. | n.d. | n.d. | n.d. |
| P17 | F | In their 20s | n.d. | n.d. | n.d. | n.d. | n.d. | n.d. | n.d. | n.d. | n.d. |
| P18 | M | In their 30s | n.d. | n.d. | n.d. | n.d. | n.d. | n.d. | n.d. | n.d. | n.d. |
| P19 | F | In their 60s | n.d. | n.d. | n.d. | n.d. | n.d. | n.d. | n.d. | n.d. | n.d. |
| P20 | F | In their 20s | n.d. | n.d. | n.d. | n.d. | n.d. | n.d. | n.d. | n.d. | n.d. |
