## Supplementary Table 9 for "Lasting first impression: Pre-existing immunity restricts mucosal antibody responses during Omicron breakthrough"

| Donor: | Age: | Gender: | Vaccination  History: | Time from last vaccination  till breakthrough  infection (months): | Timepoints post  onset for plasma /  saliva samples  (bold: incl. nasal swabs): | Breakthrough  VoC |
| --- | --- | --- | --- | --- | --- | --- |
| BT01 | In their 50s | F | 2 x BNT162b2 | 5.3 | 1, **3, 6,** 8, 16 | Delta |
| BT02 | In their 30s | M | 2 x BNT162b2 | 4.3 | **2, 6,** 8, 13, 21 | Delta |
| BT03 | In their 50s | F | 2 x ChAdOx nCoV-19 | 3 | **4, 6, 8, 12,** 16, 18, 23, 31 | Delta |
| BT04 | In their teens | F | 2 x BNT162b2 | 0.3 | 6, **15** | Delta |
| BT05 | In their 20s | M | 2 x ChAdOx nCoV-19 | 1 | 3, 10, **12,** 15, 18, 39 | Delta |
| BT06 | In their 30s | F | 2 x BNT162b2 | 4.6 | 0, **1, 2, 3, 4, 6, 7, 9,** 11, 13, 30 | Delta |
| BT07 | In their 30s | F | 2 x BNT162b2 | 3.6 | **2, 3, 4, 6, 7, 9 ,11, 13**, 30 | Delta |
| BT08 | In their 20s | M | 2 x BNT162b2 | 4.3 | **3, 4, 6, 8,** 10, 20, 27 | Delta |
| BT20 | In their 50s | F | 2 x ChAdOx nCoV-19  1 x mRNA-1273 | 4.3 | **2, 4, 6, 8,** 13, 34 | Omicron BA.2 |
| BT21 | In their 50s | M | 2 x ChAdOx nCoV-19  1 x mRNA-1273 | 4 | **5, 7, 9, 11,** 16, 37 | Omicron BA.2 |
| BT22 | In their 50s | F | 2 x ChAdOx nCoV-19  1 x mRNA-1273 | 5.3 | 2, **4, 8, 10**, 12, 38 | Omicron BA.2 |
| BT23 | In their 60s | M | 2 x ChAdOx nCoV-19  1 x mRNA-1273 | 2.6 | 0, **2, 6, 8,** 10, 36 | Omicron BA.2 |
| BT24 | In their 40s | F | 3 x BNT162b2 | 4 | **1, 2, 4**, 8, **11,** 16, 29 | Omicron BA.2 |
| BT25 | In their 50s | F | 2 x ChAdOx nCoV-19  1 x mRNA-1273 | 2 | **2,** 6, 8, **10**, 14, 35 | Omicron BA.2 |
| BT26 | In their 40s | M | 3 x BNT162b2 | 1.6 | **2, 3,** 5, 9, **12**, 17, 30 | Omicron BA.2 |
| BT27 | In their 60s | M | 2 x ChAdOx nCoV-19  1 x mRNA-1273 | 3.3 | **5,** 7, **9,** 13, 35 | Omicron BA.2 |
| BT28 | In their 40s | F | 2 x BNT162b2  1 x mRNA-1273 | 3 | **3, 6,** 10, 12, 34 | Omicron BA.2 |
| BT29 | In their 60s | M | 2 x ChAdOx nCoV-19  1 x BNT162b2 | 1.6 | **5**, 6, **8,** 12, 34 | Omicron BA.2 |

Supplementary Table 9. Cohort of SARS-CoV-2 breakthrough infections
