## Supplementary Table 10 for "Lasting first impression: Pre-existing immunity restricts mucosal antibody responses during Omicron breakthrough"

Supplementary Table 10: Parameters and confidence intervals for modelled kinetic curves of COVID-19 breakthrough cohorts

**Delta breakthrough cohort (Plasma)**

| **Sample: Plasma**  **Detector: Pan IgG** | **WT Spike 1** | | | **Delta Spike 1** | | | **BA1 Spike 1** | | | **BA2 Spike 1** | | |
| --- | --- | --- | --- | --- | --- | --- | --- | --- | --- | --- | --- | --- |
|  | Lower CI | Mean | Upper CI | Lower CI | Mean | Upper CI | Lower CI | Mean | Upper CI | Lower CI | Mean | Upper CI |
| Initial MFI: | 2452.00 | 4570.88 | 8520.79 | 1450.37 | 2691.53 | 4994.82 | 147.64 | 416.87 | 1177.06 | 474.68 | 977.24 | 2011.87 |
| Growth rate: | 0.18 | 0.26 | 0.37 | 0.24 | 0.32 | 0.43 | 0.35 | 0.49 | 0.68 | 0.35 | 0.46 | 0.60 |
| Activation time (days): | 3.25 | 4.31 | 5.70 | 3.37 | 4.26 | 5.39 | 3.72 | 4.48 | 5.40 | 3.67 | 4.44 | 5.37 |
| Peak time (days): | 7.35 | 8.58 | 10.03 | 7.08 | 8.17 | 9.42 | 6.40 | 7.39 | 8.54 | 6.46 | 7.39 | 8.45 |
| Maximum MFI: | 60022.75 | 80786.00 | 108731.74 | 40682.38 | 58140.08 | 83089.26 | 7549.81 | 12057.31 | 19255.94 | 15847.52 | 25655.55 | 41533.76 |
| **Sample: Plasma**  **Detector: Pan IgG** | **WT Spike Trimer** | | | **Delta Spike Trimer** | | | **BA1 Spike Trimer** | | | **BA2 Spike Trimer** | | |
|  | Lower CI | Mean | Upper CI | Lower CI | Mean | Upper CI | Lower CI | Mean | Upper CI | Lower CI | Mean | Upper CI |
| Initial MFI: | 3533.13 | 7079.46 | 14185.35 | 2340.78 | 5370.32 | 12320.83 | 731.88 | 1513.56 | 3130.11 | 426.30 | 812.83 | 1549.82 |
| Growth rate: | 0.22 | 0.28 | 0.37 | 0.13 | 0.20 | 0.30 | 0.24 | 0.31 | 0.41 | 0.27 | 0.35 | 0.47 |
| Activation time (days): | 3.79 | 4.90 | 6.34 | 3.03 | 4.44 | 6.50 | 3.69 | 4.66 | 5.90 | 3.79 | 4.71 | 5.86 |
| Peak time (days): | 7.27 | 8.41 | 9.74 | 8.74 | 9.68 | 10.72 | 7.05 | 8.17 | 9.46 | 7.04 | 8.08 | 9.28 |
| Maximum MFI: | 63778.05 | 87397.54 | 119764.24 | 52597.18 | 75669.39 | 108862.42 | 19256.18 | 29932.47 | 46528.08 | 11581.74 | 19694.09 | 33488.68 |
| **Sample: Plasma**  **Detector: IgA** | **WT Spike 1** | | | **Delta Spike 1** | | | **BA1 Spike 1** | | | **BA2 Spike 1** | | |
|  | Lower CI | Mean | Upper CI | Lower CI | Mean | Upper CI | Lower CI | Mean | Upper CI | Lower CI | Mean | Upper CI |
| Initial MFI: | 29.96 | 50.12 | 83.84 | 11.71 | 19.95 | 33.98 | 10.47 | 10.47 | 10.47 | 14.45 | 14.45 | 14.45 |
| Growth rate: | 0.26 | 0.31 | 0.36 | 0.28 | 0.46 | 0.76 | 0.21 | 0.48 | 1.09 | 0.17 | 0.55 | 1.80 |
| Activation time (days): | 2.69 | 4.06 | 6.12 | 3.08 | 4.18 | 5.66 | 3.77 | 4.85 | 6.25 | 2.60 | 3.82 | 5.61 |
| Peak time (days): | 7.50 | 8.94 | 10.65 | 6.51 | 7.69 | 9.09 | 6.44 | 7.32 | 8.31 | 5.69 | 7.17 | 9.04 |
| Maximum MFI: | 1139.05 | 3602.69 | 11394.95 | 731.38 | 2240.54 | 6863.79 | 59.48 | 202.24 | 687.63 | 144.51 | 506.60 | 1776.00 |
| **Sample: Plasma**  **Detector: IgA** | **WT Spike Trimer** | | | **Delta Spike Trimer** | | | **BA1 Spike Trimer** | | | **BA2 Spike Trimer** | | |
|  | Lower CI | Mean | Upper CI | Lower CI | Mean | Upper CI | Lower CI | Mean | Upper CI | Lower CI | Mean | Upper CI |
| Initial MFI: | 40.39 | 102.33 | 259.27 | 27.75 | 79.43 | 227.34 | 17.38 | 34.67 | 69.16 | 16.98 | 16.98 | 16.98 |
| Growth rate: | 0.26 | 0.32 | 0.40 | 0.27 | 0.32 | 0.38 | 0.21 | 0.32 | 0.48 | 0.19 | 0.33 | 0.57 |
| Activation time (days): | 2.97 | 4.01 | 5.43 | 2.63 | 3.60 | 4.91 | 2.52 | 3.71 | 5.44 | 2.51 | 3.78 | 5.70 |
| Peak time (days): | 7.08 | 8.50 | 10.21 | 6.83 | 8.33 | 10.15 | 6.93 | 8.08 | 9.44 | 7.06 | 8.25 | 9.64 |
| Maximum MFI: | 2715.44 | 6904.71 | 17557.04 | 2572.90 | 6526.89 | 16557.30 | 784.21 | 2646.87 | 8933.70 | 422.03 | 1412.80 | 4729.51 |
| **Sample: Plasma**  **Detector: FcγR3a** | **WT Spike 1** | | | **Delta Spike 1** | | | **BA1 Spike 1** | | | **BA2 Spike 1** | | |
|  | Lower CI | Mean | Upper CI | Lower CI | Mean | Upper CI | Lower CI | Mean | Upper CI | Lower CI | Mean | Upper CI |
| Initial MFI: | 10.58 | 22.39 | 47.35 | 7.73 | 7.73 | 7.73 | 7.00 | 7.00 | 7.00 | 12.02 | 12.02 | 12.02 |
| Growth rate: | 0.29 | 0.41 | 0.58 | 0.39 | 0.63 | 1.02 | 0.29 | 0.71 | 1.74 | 0.32 | 0.72 | 1.59 |
| Activation time (days): | 3.04 | 4.14 | 5.63 | 3.18 | 4.26 | 5.72 | 4.23 | 5.16 | 6.28 | 3.69 | 4.62 | 5.77 |
| Peak time (days): | 7.43 | 8.58 | 9.92 | 6.88 | 7.85 | 8.95 | 6.13 | 6.89 | 7.74 | 6.13 | 6.89 | 7.75 |
| Maximum MFI: | 1149.22 | 2440.14 | 5181.15 | 500.78 | 1307.45 | 3413.48 | 34.33 | 81.56 | 193.77 | 80.78 | 289.76 | 1039.33 |
| **Sample: Plasma**  **Detector: FcγR3a** | **WT Spike Trimer** | | | **Delta Spike Trimer** | | | **BA1 Spike Trimer** | | | **BA2 Spike Trimer** | | |
|  | Lower CI | Mean | Upper CI | Lower CI | Mean | Upper CI | Lower CI | Mean | Upper CI | Lower CI | Mean | Upper CI |
| Initial MFI: | 19.93 | 58.88 | 173.94 | 15.26 | 35.48 | 82.51 | 41.69 | 41.69 | 41.69 | 30.20 | 30.20 | 30.20 |
| Growth rate: | 0.14 | 0.24 | 0.39 | 0.22 | 0.33 | 0.50 | 0.12 | 0.22 | 0.40 | 0.17 | 0.28 | 0.46 |
| Activation time (days): | 3.06 | 3.90 | 4.97 | 3.01 | 4.35 | 6.29 | 4.99 | 5.99 | 7.19 | 4.11 | 4.90 | 5.85 |
| Peak time (days): | 7.97 | 9.49 | 11.29 | 7.96 | 9.03 | 10.23 | 9.77 | 11.36 | 13.21 | 9.28 | 10.70 | 12.34 |
| Maximum MFI: | 1280.79 | 2880.93 | 6480.22 | 889.36 | 2214.45 | 5513.83 | 206.32 | 569.38 | 1571.34 | 116.46 | 360.96 | 1118.75 |

**Delta breakthrough cohort (Saliva)**

| **Sample: Saliva**  **Detector: Pan IgG** | **WT Spike 1** | | | **Delta Spike 1** | | | **BA1 Spike 1** | | | **BA2 Spike 1** | | |
| --- | --- | --- | --- | --- | --- | --- | --- | --- | --- | --- | --- | --- |
|  | Lower CI | Mean | Upper CI | Lower CI | Mean | Upper CI | Lower CI | Mean | Upper CI | Lower CI | Mean | Upper CI |
| Initial MFI: | 1456.26 | 2630.27 | 4750.73 | 757.74 | 1412.54 | 2633.18 | 83.96 | 208.93 | 519.90 | 196.01 | 398.11 | 808.57 |
| Growth rate: | 0.23 | 0.28 | 0.33 | 0.15 | 0.32 | 0.67 | 0.26 | 0.36 | 0.51 | 0.18 | 0.40 | 0.90 |
| Activation time (days): | 3.24 | 4.81 | 7.14 | 3.21 | 4.81 | 7.20 | 2.03 | 4.18 | 8.61 | 2.03 | 4.39 | 9.51 |
| Peak time (days): | 8.61 | 10.07 | 11.79 | 6.98 | 9.87 | 13.97 | 7.72 | 9.12 | 10.77 | 7.36 | 8.94 | 10.85 |
| Maximum MFI: | 52419.76 | 82960.99 | 131296.40 | 34349.48 | 59621.86 | 103488.23 | 8189.38 | 15292.07 | 28554.98 | 15572.19 | 29617.63 | 56331.43 |
| **Sample: Saliva**  **Detector: Pan IgG** | **WT Spike Trimer** | | | **Delta Spike Trimer** | | | **BA1 Spike Trimer** | | | **BA2 Spike Trimer** | | |
|  | Lower CI | Mean | Upper CI | Lower CI | Mean | Upper CI | Lower CI | Mean | Upper CI | Lower CI | Mean | Upper CI |
| Initial MFI: | 2333.24 | 4570.88 | 8954.47 | 1778.77 | 3548.13 | 7077.50 | 791.63 | 1318.26 | 2195.23 | 453.48 | 758.58 | 1268.94 |
| Growth rate: | 0.16 | 0.24 | 0.37 | 0.18 | 0.26 | 0.39 | 0.18 | 0.26 | 0.38 | 0.22 | 0.28 | 0.35 |
| Activation time (days): | 3.56 | 5.10 | 7.32 | 3.58 | 5.10 | 7.28 | 3.58 | 5.21 | 7.57 | 3.74 | 5.05 | 6.82 |
| Peak time (days): | 8.32 | 10.38 | 12.95 | 8.49 | 10.18 | 12.19 | 8.30 | 10.18 | 12.48 | 8.74 | 10.07 | 11.61 |
| Maximum MFI: | 59092.74 | 83926.20 | 119195.80 | 48672.35 | 72419.92 | 107754.08 | 19554.12 | 31559.85 | 50936.77 | 11795.82 | 21236.87 | 38234.29 |
| **Sample: Saliva**  **Detector: IgA** | **WT Spike 1** | | | **Delta Spike 1** | | | **BA1 Spike 1** | | | **BA2 Spike 1** | | |
|  | Lower CI | Mean | Upper CI | Lower CI | Mean | Upper CI | Lower CI | Mean | Upper CI | Lower CI | Mean | Upper CI |
| Initial MFI: | 1277.50 | 2089.30 | 3416.96 | 586.81 | 870.96 | 1292.71 | 12.02 | 12.02 | 12.02 | 7.80 | 19.05 | 46.57 |
| Growth rate: | 0.18 | 0.32 | 0.56 | 0.13 | 0.28 | 0.60 | 0.21 | 0.32 | 0.49 | 0.20 | 0.35 | 0.62 |
| Activation time (days): | 6.41 | 7.39 | 8.52 | 5.64 | 7.10 | 8.93 | 4.55 | 6.05 | 8.04 | 3.78 | 5.58 | 8.25 |
| Peak time (days): | 8.50 | 9.49 | 10.59 | 8.76 | 9.97 | 11.35 | 8.27 | 9.12 | 10.05 | 8.50 | 10.38 | 12.68 |
| Maximum MFI: | 12793.77 | 24412.07 | 46581.21 | 24012.78 | 37021.41 | 57077.32 | 5170.17 | 8866.42 | 15205.17 | 6382.85 | 11109.55 | 19336.50 |
| **Sample: Saliva**  **Detector: IgA** | **WT Spike Trimer** | | | **Delta Spike Trimer** | | | **BA1 Spike Trimer** | | | **BA2 Spike Trimer** | | |
|  | Lower CI | Mean | Upper CI | Lower CI | Mean | Upper CI | Lower CI | Mean | Upper CI | Lower CI | Mean | Upper CI |
| Initial MFI: | 405.70 | 1023.29 | 2581.07 | 354.29 | 812.83 | 1864.83 | 2699.48 | 4897.79 | 8886.28 | 1587.32 | 2454.71 | 3796.09 |
| Growth rate: | 0.13 | 0.22 | 0.39 | 0.13 | 0.24 | 0.43 | 0.08 | 0.20 | 0.47 | 0.02 | 0.13 | 0.99 |
| Activation time (days): | 3.77 | 5.64 | 8.43 | 3.50 | 5.21 | 7.75 | 7.86 | 9.30 | 11.00 | 6.89 | 9.49 | 13.06 |
| Peak time (days): | 9.03 | 10.70 | 12.67 | 8.54 | 10.28 | 12.37 | 9.55 | 11.25 | 13.24 | 8.96 | 12.43 | 17.24 |
| Maximum MFI: | 10434.39 | 26618.42 | 67904.31 | 44129.44 | 60879.45 | 83987.18 | 12923.19 | 22500.64 | 39176.01 | 6465.54 | 12044.92 | 22438.96 |
| **Sample: Saliva**  **Detector: FcγR3a** | **WT Spike 1** | | | **Delta Spike 1** | | | **BA1 Spike 1** | | | **BA2 Spike 1** | | |
|  | Lower CI | Mean | Upper CI | Lower CI | Mean | Upper CI | Lower CI | Mean | Upper CI | Lower CI | Mean | Upper CI |
| Initial MFI: | 6.03 | 15.49 | 39.78 | 18.20 | 18.20 | 18.20 | 16.98 | 16.98 | 16.98 | 11.75 | 11.75 | 11.75 |
| Growth rate: | 0.26 | 0.39 | 0.60 | 0.22 | 0.35 | 0.54 | 0.12 | 0.23 | 0.47 | 0.27 | 0.40 | 0.59 |
| Activation time (days): | 2.94 | 4.53 | 6.98 | 3.66 | 5.10 | 7.11 | 4.72 | 6.05 | 7.76 | 4.57 | 5.81 | 7.40 |
| Peak time (days): | 8.51 | 10.07 | 11.92 | 9.43 | 10.91 | 12.63 | 6.88 | 8.50 | 10.50 | 7.67 | 9.03 | 10.61 |
| Maximum MFI: | 1045.81 | 2594.15 | 6434.81 | 446.74 | 1508.93 | 5096.59 | 26.45 | 111.68 | 471.51 | 79.56 | 394.54 | 1956.52 |
| **Sample: Saliva**  **Detector: FcγR3a** | **WT Spike Trimer** | | | **Delta Spike Trimer** | | | **BA1 Spike Trimer** | | | **BA2 Spike Trimer** | | |
|  | Lower CI | Mean | Upper CI | Lower CI | Mean | Upper CI | Lower CI | Mean | Upper CI | Lower CI | Mean | Upper CI |
| Initial MFI: | 18.53 | 44.67 | 107.70 | 11.60 | 23.99 | 49.61 | 23.99 | 23.99 | 23.99 | 20.89 | 20.89 | 20.89 |
| Growth rate: | 0.16 | 0.25 | 0.39 | 0.21 | 0.30 | 0.44 | 0.13 | 0.24 | 0.44 | 0.17 | 0.30 | 0.52 |
| Activation time (days): | 3.11 | 4.90 | 7.73 | 3.64 | 5.21 | 7.44 | 3.72 | 5.37 | 7.74 | 2.15 | 3.63 | 6.13 |
| Peak time (days): | 9.80 | 11.70 | 13.97 | 9.80 | 11.36 | 13.17 | 9.12 | 11.13 | 13.60 | 8.62 | 10.59 | 13.01 |
| Maximum MFI: | 1705.03 | 3455.75 | 7004.10 | 1193.14 | 2761.96 | 6393.56 | 230.85 | 707.85 | 2170.46 | 146.19 | 490.87 | 1648.24 |

**Omicron BA.2 breakthrough cohort (Plasma)**

| **Sample: Plasma**  **Detector: Pan IgG** | **WT Spike 1** | | | **Delta Spike 1** | | | **BA1 Spike 1** | | | **BA2 Spike 1** | | |
| --- | --- | --- | --- | --- | --- | --- | --- | --- | --- | --- | --- | --- |
|  | Lower CI | Mean | Upper CI | Lower CI | Mean | Upper CI | Lower CI | Mean | Upper CI | Lower CI | Mean | Upper CI |
| Initial MFI: | 13688.00 | 19952.62 | 29084.39 | 6957.29 | 11481.54 | 18947.85 | 921.38 | 1905.46 | 3940.58 | 2505.65 | 4365.16 | 7604.66 |
| Growth rate: | 0.04 | 0.06 | 0.08 | 0.05 | 0.07 | 0.10 | 0.14 | 0.29 | 0.58 | 0.05 | 0.07 | 0.11 |
| Activation time (days): | 6.89 | 8.41 | 10.28 | 6.30 | 7.92 | 9.97 | 6.57 | 7.39 | 8.32 | 4.87 | 6.17 | 7.82 |
| Peak time (days): | 14.68 | 16.78 | 19.17 | 12.81 | 14.59 | 16.61 | 8.06 | 8.76 | 9.51 | 11.89 | 14.01 | 16.52 |
| Maximum MFI: | 43394.17 | 64444.81 | 95707.19 | 24012.78 | 37021.41 | 57077.32 | 3585.41 | 5948.38 | 9868.69 | 6382.85 | 11109.55 | 19336.50 |
| **Sample: Plasma**  **Detector: Pan IgG** | **WT Spike Trimer** | | | **Delta Spike Trimer** | | | **BA1 Spike Trimer** | | | **BA2 Spike Trimer** | | |
|  | Lower CI | Mean | Upper CI | Lower CI | Mean | Upper CI | Lower CI | Mean | Upper CI | Lower CI | Mean | Upper CI |
| Initial MFI: | 23722.26 | 33113.11 | 46221.50 | 12495.68 | 21379.62 | 36579.69 | 6604.93 | 9549.93 | 13808.04 | 3712.62 | 5623.41 | 8517.65 |
| Growth rate: | 0.04 | 0.07 | 0.13 | 0.02 | 0.03 | 0.05 | 0.04 | 0.06 | 0.08 | 0.04 | 0.06 | 0.09 |
| Activation time (days): | 7.81 | 9.39 | 11.30 | 4.49 | 5.87 | 7.68 | 6.46 | 7.92 | 9.72 | 5.49 | 6.89 | 8.65 |
| Peak time (days): | 12.57 | 14.15 | 15.94 | 14.84 | 17.81 | 21.38 | 14.57 | 16.78 | 19.32 | 14.78 | 16.95 | 19.43 |
| Maximum MFI: | 58368.56 | 79623.49 | 108618.41 | 44129.44 | 60879.45 | 83987.18 | 21860.87 | 34326.63 | 53900.75 | 17467.57 | 25411.93 | 36969.44 |
| **Sample: Plasma**  **Detector: IgA** | **WT Spike 1** | | | **Delta Spike 1** | | | **BA1 Spike 1** | | | **BA2 Spike 1** | | |
|  | Lower CI | Mean | Upper CI | Lower CI | Mean | Upper CI | Lower CI | Mean | Upper CI | Lower CI | Mean | Upper CI |
| Initial MFI: | 68.30 | 141.25 | 292.12 | 21.08 | 40.74 | 78.73 | 10.47 | 10.47 | 10.47 | 14.45 | 14.45 | 14.45 |
| Growth rate: | 0.07 | 0.10 | 0.16 | 0.07 | 0.12 | 0.20 | 0.11 | 0.35 | 1.08 | 0.08 | 0.12 | 0.18 |
| Activation time (days): | 4.59 | 6.17 | 8.30 | 3.23 | 4.66 | 6.73 | 6.10 | 8.00 | 10.51 | 1.73 | 2.97 | 5.12 |
| Peak time (days): | 14.12 | 16.95 | 20.34 | 12.07 | 15.49 | 19.86 | 10.42 | 12.68 | 15.43 | 13.81 | 16.78 | 20.39 |
| Maximum MFI: | 1214.34 | 2706.92 | 6034.04 | 573.14 | 1200.10 | 2512.88 | 40.95 | 103.61 | 262.15 | 213.79 | 529.71 | 1312.47 |
| **Sample: Plasma**  **Detector: IgA** | **WT Spike Trimer** | | | **Delta Spike Trimer** | | | **BA1 Spike Trimer** | | | **BA2 Spike Trimer** | | |
|  | Lower CI | Mean | Upper CI | Lower CI | Mean | Upper CI | Lower CI | Mean | Upper CI | Lower CI | Mean | Upper CI |
| Initial MFI: | 165.55 | 288.40 | 502.44 | 137.57 | 229.09 | 381.49 | 56.43 | 104.71 | 194.32 | 49.52 | 85.11 | 146.29 |
| Growth rate: | 0.04 | 0.07 | 0.12 | 0.04 | 0.07 | 0.13 | 0.05 | 0.07 | 0.11 | 0.10 | 0.15 | 0.20 |
| Activation time (days): | 3.88 | 5.31 | 7.27 | 3.65 | 5.16 | 7.28 | 2.59 | 4.22 | 6.88 | 6.55 | 8.33 | 10.60 |
| Peak time (days): | 14.93 | 19.30 | 24.95 | 14.17 | 18.36 | 23.78 | 14.27 | 17.64 | 21.79 | 13.25 | 15.33 | 17.74 |
| Maximum MFI: | 1876.28 | 4043.89 | 8715.69 | 1485.65 | 3058.28 | 6295.62 | 671.78 | 1487.06 | 3291.78 | 381.20 | 908.36 | 2164.53 |
| **Sample: Plasma**  **Detector: FcγR3a** | **WT Spike 1** | | | **Delta Spike 1** | | | **BA1 Spike 1** | | | **BA2 Spike 1** | | |
|  | Lower CI | Mean | Upper CI | Lower CI | Mean | Upper CI | Lower CI | Mean | Upper CI | Lower CI | Mean | Upper CI |
| Initial MFI: | 106.65 | 208.93 | 409.30 | 45.39 | 79.43 | 139.01 | 7.00 | 7.00 | 7.00 | 8.52 | 14.13 | 23.42 |
| Growth rate: | 0.04 | 0.07 | 0.11 | 0.04 | 0.06 | 0.09 | 0.05 | 0.11 | 0.27 | 0.06 | 0.09 | 0.14 |
| Activation time (days): | 5.86 | 7.32 | 9.13 | 4.60 | 5.99 | 7.80 | 6.10 | 8.00 | 10.51 | 2.41 | 4.14 | 7.11 |
| Peak time (days): | 15.88 | 18.92 | 22.53 | 15.50 | 18.73 | 22.62 | 8.98 | 11.36 | 14.37 | 13.53 | 16.95 | 21.23 |
| Maximum MFI: | 728.37 | 1670.04 | 3829.10 | 258.17 | 629.14 | 1533.16 | 11.66 | 21.37 | 39.15 | 92.16 | 216.25 | 507.43 |
| **Sample: Plasma**  **Detector: FcγR3a** | **WT Spike Trimer** | | | **Delta Spike Trimer** | | | **BA1 Spike Trimer** | | | **BA2 Spike Trimer** | | |
|  | Lower CI | Mean | Upper CI | Lower CI | Mean | Upper CI | Lower CI | Mean | Upper CI | Lower CI | Mean | Upper CI |
| Initial MFI: | 283.19 | 562.34 | 1116.66 | 149.10 | 346.74 | 806.34 | 62.96 | 91.20 | 132.10 | 36.94 | 53.70 | 78.07 |
| Growth rate: | 0.03 | 0.05 | 0.09 | 0.03 | 0.05 | 0.09 | 0.04 | 0.07 | 0.11 | 0.08 | 0.12 | 0.18 |
| Activation time (days): | 5.71 | 7.10 | 8.82 | 4.88 | 6.42 | 8.45 | 6.06 | 7.54 | 9.37 | 6.73 | 8.50 | 10.73 |
| Peak time (days): | 14.89 | 17.81 | 21.32 | 14.45 | 17.46 | 21.11 | 15.59 | 18.92 | 22.95 | 13.47 | 16.12 | 19.29 |
| Maximum MFI: | 1514.36 | 2750.59 | 4996.00 | 974.73 | 1835.16 | 3455.11 | 279.84 | 713.45 | 1818.96 | 190.99 | 446.85 | 1045.46 |

**Omicron BA.2 breakthrough cohort (Saliva)**

| **Sample: Saliva**  **Detector: Pan IgG** | **WT Spike 1** | | | **Delta Spike 1** | | | **BA1 Spike 1** | | | **BA2 Spike 1** | | |
| --- | --- | --- | --- | --- | --- | --- | --- | --- | --- | --- | --- | --- |
|  | Lower CI | Mean | Upper CI | Lower CI | Mean | Upper CI | Lower CI | Mean | Upper CI | Lower CI | Mean | Upper CI |
| Initial MFI: | 6322.37 | 10964.78 | 19016.04 | 2778.69 | 5370.32 | 10379.11 | 543.75 | 1000.00 | 1839.08 | 1040.78 | 1905.46 | 3488.51 |
| Growth rate: | 0.01 | 0.27 | 6.03 | 0.00 | 0.27 | 17.04 | 0.00 | 0.27 | 454.14 | 0.02 | 0.25 | 2.78 |
| Activation time (days): | 4.84 | 7.17 | 10.63 | 4.06 | 7.10 | 12.41 | 3.11 | 7.24 | 16.86 | 4.76 | 7.24 | 11.02 |
| Peak time (days): | 6.87 | 8.33 | 10.10 | 6.66 | 8.25 | 10.21 | 6.44 | 8.17 | 10.35 | 7.26 | 9.30 | 11.91 |
| Maximum MFI: | 26945.80 | 45975.98 | 78446.02 | 12113.07 | 24084.05 | 47885.57 | 2331.57 | 4606.06 | 9099.35 | 6490.79 | 12285.01 | 23251.65 |
| **Sample: Saliva**  **Detector: Pan IgG** | **WT Spike Trimer** | | | **Delta Spike Trimer** | | | **BA1 Spike Trimer** | | | **BA2 Spike Trimer** | | |
|  | Lower CI | Mean | Upper CI | Lower CI | Mean | Upper CI | Lower CI | Mean | Upper CI | Lower CI | Mean | Upper CI |
| Initial MFI: | 12478.43 | 19952.62 | 31903.62 | 7999.08 | 14125.38 | 24943.65 | 4326.69 | 6760.83 | 10564.38 | 1963.54 | 3467.37 | 6122.94 |
| Growth rate: | 0.02 | 0.19 | 2.02 | 0.00 | 0.22 | 1071.06 | 0.03 | 0.23 | 1.65 | 0.02 | 0.19 | 1.88 |
| Activation time (days): | 5.17 | 6.75 | 8.82 | 4.98 | 6.82 | 9.35 | 5.48 | 6.89 | 8.67 | 4.58 | 7.17 | 11.23 |
| Peak time (days): | 6.07 | 7.92 | 10.35 | 2.77 | 7.92 | 22.66 | 6.54 | 8.00 | 9.80 | 7.17 | 9.39 | 12.31 |
| Maximum MFI: | 39325.96 | 62988.29 | 100888.18 | 29573.63 | 50590.79 | 86544.28 | 17800.46 | 28637.84 | 46073.28 | 9207.57 | 17454.63 | 33088.45 |
| **Sample: Saliva**  **Detector: IgA** | **WT Spike 1** | | | **Delta Spike 1** | | | **BA1 Spike 1** | | | **BA2 Spike 1** | | |
|  | Lower CI | Mean | Upper CI | Lower CI | Mean | Upper CI | Lower CI | Mean | Upper CI | Lower CI | Mean | Upper CI |
| Initial MFI: | 1031.43 | 1905.46 | 3520.14 | 393.59 | 870.96 | 1927.35 | 12.02 | 12.02 | 12.02 | 1.31 | 5.42 | 22.46 |
| Growth rate: | 0.05 | 0.10 | 0.22 | 0.06 | 0.13 | 0.26 | 0.19 | 0.33 | 0.57 | 0.18 | 0.29 | 0.47 |
| Activation time (days): | 6.70 | 8.85 | 11.69 | 6.51 | 8.33 | 10.66 | 8.79 | 11.47 | 14.98 | 3.49 | 5.37 | 8.26 |
| Peak time (days): | 11.50 | 14.01 | 17.08 | 10.28 | 12.55 | 15.33 | 12.29 | 15.33 | 19.13 | 10.95 | 13.20 | 15.90 |
| Maximum MFI: | 5968.77 | 12172.47 | 24824.05 | 2921.63 | 6310.56 | 13630.49 | 151.31 | 310.02 | 635.21 | 745.41 | 2012.94 | 5435.84 |
| **Sample: Saliva**  **Detector: IgA** | **WT Spike Trimer** | | | **Delta Spike Trimer** | | | **BA1 Spike Trimer** | | | **BA2 Spike Trimer** | | |
|  | Lower CI | Mean | Upper CI | Lower CI | Mean | Upper CI | Lower CI | Mean | Upper CI | Lower CI | Mean | Upper CI |
| Initial MFI: | 666.56 | 1288.25 | 2489.77 | 417.37 | 870.96 | 1817.52 | 2197.05 | 3388.44 | 5225.89 | 1019.76 | 1513.56 | 2246.47 |
| Growth rate: | 0.09 | 0.17 | 0.31 | 0.10 | 0.16 | 0.25 | 0.05 | 0.15 | 0.46 | 0.05 | 0.18 | 0.67 |
| Activation time (days): | 6.06 | 8.08 | 10.78 | 5.63 | 7.39 | 9.70 | 2.88 | 4.44 | 6.83 | 4.43 | 6.42 | 9.32 |
| Peak time (days): | 11.60 | 13.74 | 16.26 | 11.75 | 14.15 | 17.05 | 4.90 | 6.82 | 9.50 | 7.25 | 9.30 | 11.93 |
| Maximum MFI: | 11746.97 | 18413.92 | 28864.67 | 10935.70 | 16786.36 | 25767.15 | 13049.94 | 17791.50 | 24255.84 | 7598.80 | 10574.82 | 14716.37 |
| **Sample: Saliva**  **Detector: FcγR3a** | **WT Spike 1** | | | **Delta Spike 1** | | | **BA1 Spike 1** | | | **BA2 Spike 1** | | |
|  | Lower CI | Mean | Upper CI | Lower CI | Mean | Upper CI | Lower CI | Mean | Upper CI | Lower CI | Mean | Upper CI |
| Initial MFI: | 81.46 | 177.83 | 388.22 | 20.32 | 50.12 | 123.59 | 16.98 | 16.98 | 16.98 | 10.23 | 10.23 | 10.23 |
| Growth rate: | 0.02 | 0.22 | 2.61 | 0.00 | 0.22 | 10.40 | 0.00 | 0.08 | 2.99 | 0.04 | 0.16 | 0.64 |
| Activation time (days): | 5.28 | 8.00 | 12.13 | 3.43 | 7.24 | 15.28 | 3.68 | 6.62 | 11.92 | 1.66 | 4.35 | 11.36 |
| Peak time (days): | 7.00 | 10.07 | 14.51 | 5.97 | 9.30 | 14.48 | 5.07 | 8.50 | 14.26 | 5.69 | 9.49 | 15.82 |
| Maximum MFI: | 695.15 | 1797.52 | 4647.99 | 216.73 | 663.87 | 2033.50 | 26.12 | 46.70 | 83.51 | 64.77 | 215.22 | 715.20 |
| **Sample: Saliva**  **Detector: FcγR3a** | **WT Spike Trimer** | | | **Delta Spike Trimer** | | | **BA1 Spike Trimer** | | | **BA2 Spike Trimer** | | |
|  | Lower CI | Mean | Upper CI | Lower CI | Mean | Upper CI | Lower CI | Mean | Upper CI | Lower CI | Mean | Upper CI |
| Initial MFI: | 190.44 | 457.09 | 1097.08 | 113.26 | 281.84 | 701.33 | 31.80 | 66.07 | 137.25 | 18.33 | 41.69 | 94.78 |
| Growth rate: | 0.00 | 0.20 | 3305836.17 | 0.00 | 0.14 | 8.31 | 0.03 | 0.18 | 1.07 | 0.03 | 0.15 | 0.77 |
| Activation time (days): | 0.99 | 7.17 | 51.91 | 4.16 | 7.61 | 13.95 | 3.91 | 6.05 | 9.37 | 4.03 | 6.89 | 11.79 |
| Peak time (days): | 2.78 | 8.58 | 26.55 | 5.11 | 9.97 | 19.46 | 5.55 | 8.76 | 13.83 | 7.14 | 10.70 | 16.02 |
| Maximum MFI: | 913.69 | 2515.28 | 6924.24 | 592.77 | 1823.82 | 5611.56 | 296.24 | 785.77 | 2084.23 | 180.93 | 535.25 | 1583.47 |
